# Transitions in lung microbiota landscape associate with distinct patterns of pneumonia progression

**DOI:** 10.1101/2024.08.02.24311426

**Authors:** Jack T. Sumner, Chiagozie I. Pickens, Stefanie Huttelmaier, Anahid A. Moghadam, Hiam Abdala-Valencia, NU SCRIPT Study Investigators, Alan R Hauser, Patrick C. Seed, Richard G. Wunderink, Erica M. Hartmann

**Affiliations:** Department of Civil and Environmental Engineering, Northwestern University, Evanston, IL, USA; Department of Medicine, Division of Pulmonary and Critical Care, Northwestern University, Chicago, IL, USA; Northwestern University Successful Clinical Response in Pneumonia Therapy (NU SCRIPT); Department of Microbiology-Immunology, Northwestern University, Chicago, IL, USA; Department of Medicine, Division of Pediatric Infectious Diseases, Ann & Robert H. Lurie Children’s Hospital of Chicago, Chicago, IL, USA; Center for Synthetic Biology, Northwestern University, Evanston, IL, USA

## Abstract

The precise microbial determinants driving clinical outcomes in severe pneumonia are unknown. Competing ecological forces produce dynamic microbiota states in health; infection and treatment effects on microbiota state must be defined to improve pneumonia therapy. Here, we leverage our unique clinical setting, which includes systematic and serial bronchoscopic sampling in patients with suspected pneumonia, to determine lung microbial ecosystem dynamics throughout pneumonia therapy. We combine 16S rRNA gene amplicon, metagenomic, and transcriptomic sequencing with bacterial load quantification to reveal clinically-relevant pneumonia progression drivers. Microbiota states are predictive of pneumonia category and exhibit differential stability and pneumonia therapy response. Disruptive forces, like aspiration, associate with cohesive changes in gene expression and microbial community structure. In summary, we show that host and microbiota landscapes change in unison with clinical phenotypes and that microbiota state dynamics reflect pneumonia progression. We suggest that distinct pathways of lung microbial community succession mediate pneumonia progression.

**Graphical Abstract 1.**
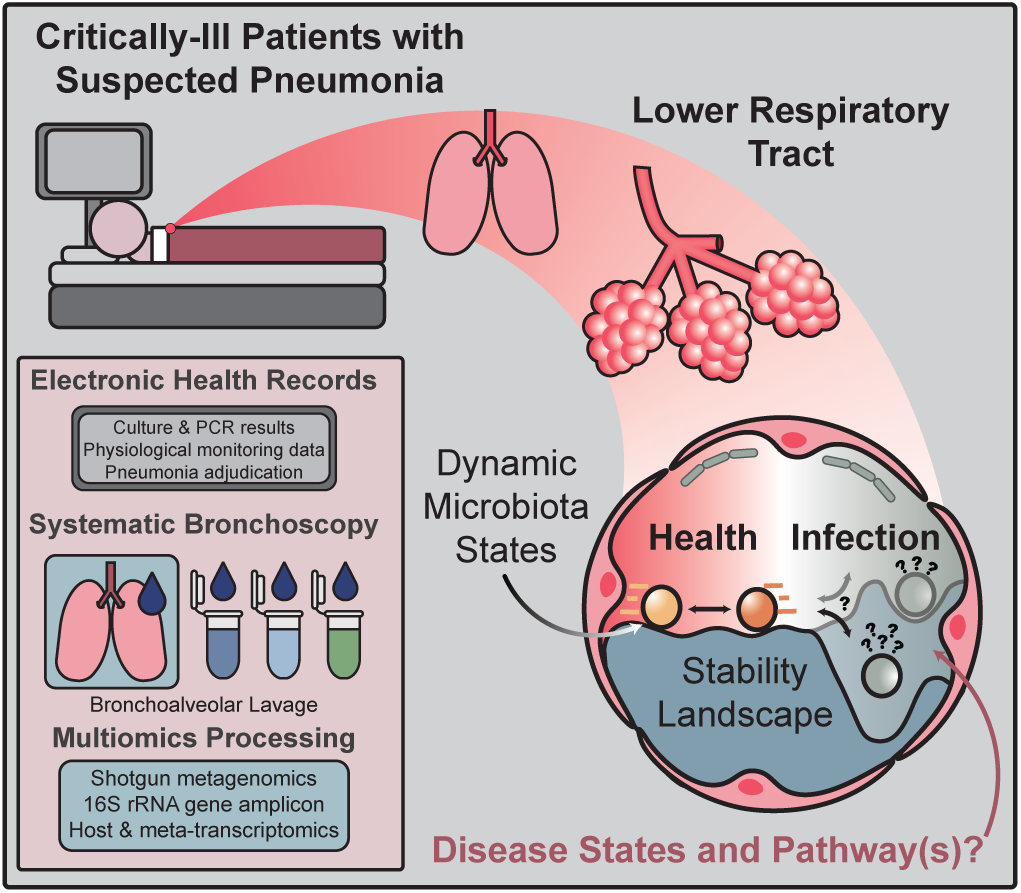
Determining landscape dynamics of lung microbial ecosystems with multiomics.

## INTRODUCTION

The classical conceptualization of pneumonia pathogenesis disregards the contribution of the normal lung microbiome^1^. A paucity of data results in poor understanding of the microbial determinants driving pneumonia outcome^2^. Three general categories of pneumonia, ventilator-associated (VAP), hospital-acquired (HAP), and community-acquired (CAP), are each associated with specific pathogens^3^. This differentiation indicates pneumonia may associate with discrete microbiota states (e.g. conserved combinations of microorganisms called pneumotypes) at the time of diagnosis. If true, it would suggest divergent community succession pathways precede microbiota state development. Similarly, the application of antimicrobials is expected to promote divergent community succession pathways, depending on initial microbiota state and successful treatment response.

Microbial colonization from other niches and clinical practices at least partially drive distal lung microbiome composition^4–7^. Early evidence suggests oral-associated microbiota states play a protective role in respiratory health, both in observational human cohort studies^8–10^ and in experimental mouse models^7,11,12^. Pneumotypes enriched with oral-associated microbiota exhibit a subclinical Th17 inflammatory phenotype, suggesting commensal airway microbiota contribute to pulmonary immune function regulation^9^. An elevated oral-associated microbiota is linked with improved lung transplant success and a reduced risk of developing HAP^8,10^. Detection of salivary amylase in bronchoalveolar lavage (BAL) is associated with a greater risk of bacterial pneumonia and positive respiratory culture, although its relationship to microbiota state is not known^13,14^. The extent to which lung microbiota confer resilience or susceptibility to pneumonia, and how this function differs between CAP and HAP or VAP subtypes, remains uncertain.

Host physiological components are hypothesized to be the major driving ecological force in microbial community assembly^1,15^. However, physiology in ICU patients is often disturbed, likely playing a role in subsequent HAP acquisition. Using the data-rich clinical setting of the ICU combined with systems biology approach, we quantify the relationship between markers of physiological disruption and changes to the nascent microbial communities. To determine the microbial signatures implicated in pneumonia pathogenesis and clinical outcome, we implement a comprehensive multiomics approach, involving systematic and serial bronchoscopic sampling of over 200 critically ill patients across various pneumonia subtypes (CAP, HAP, VAP) and non-pneumonia (NP) states. We show that lung microbiota are altered in a disease-specific manner and that state-dependent transitions in the lung microbiota landscape correlate with clinical outcome. We suggest that distinct pathways of lung microbial community succession mediate pneumonia pathogenesis.

## RESULTS

### Demographics of the cohort

Bronchoalveolar lavage samples were collected as part of the Successful Clinical Response in Pneumonia Therapy (SCRIPT) Systems Biology Center, a prospective, observational study of mechanically ventilated patients with suspected pneumonia at Northwestern Memorial Hospital. Between June 2018 to June 2020, 251 participants were enrolled in SCRIPT for whom we report at least one omics profile. A standardized protocol for physician adjudication identified 54 cases of CAP, 101 HAP, and 82 VAP episodes; 33 critically-ill patients with suspected pneumonia were adjudicated to have alternative diagnoses (Table S1). Details of the adjudication process are published elsewhere^16^. The most prevalent clinical microbiologic etiologies were bacterial pneumonia followed by viral and culture-negative pneumonia (Table S1). Of the 251 total patients, 62 underwent serial BAL sampling, resulting in 345 total BAL samples. We obtained amplicon sequencing profiles from 232 samples, shotgun sequencing profiles from 215 samples, and transriptomics profiles from 218 samples ((Figure 1A, see methods for detailed inclusion criteria). Transcriptomes are derived from alveolar macrophages and cell-associated microbiota isolated using fluorescence-activated cell sorting (FACS). An additional 30 metagenomic BAL samples and 1 transcriptomic BAL sample failed library preparation. In addition, we quantified total bacterial load using qPCR in 157 samples. Samples with less than 5 µL remaining volume were omitted from quantification. The sampling overview is available as a summary (Figure 1a) and per-patient level (Figure S1).

**Figure 1.**
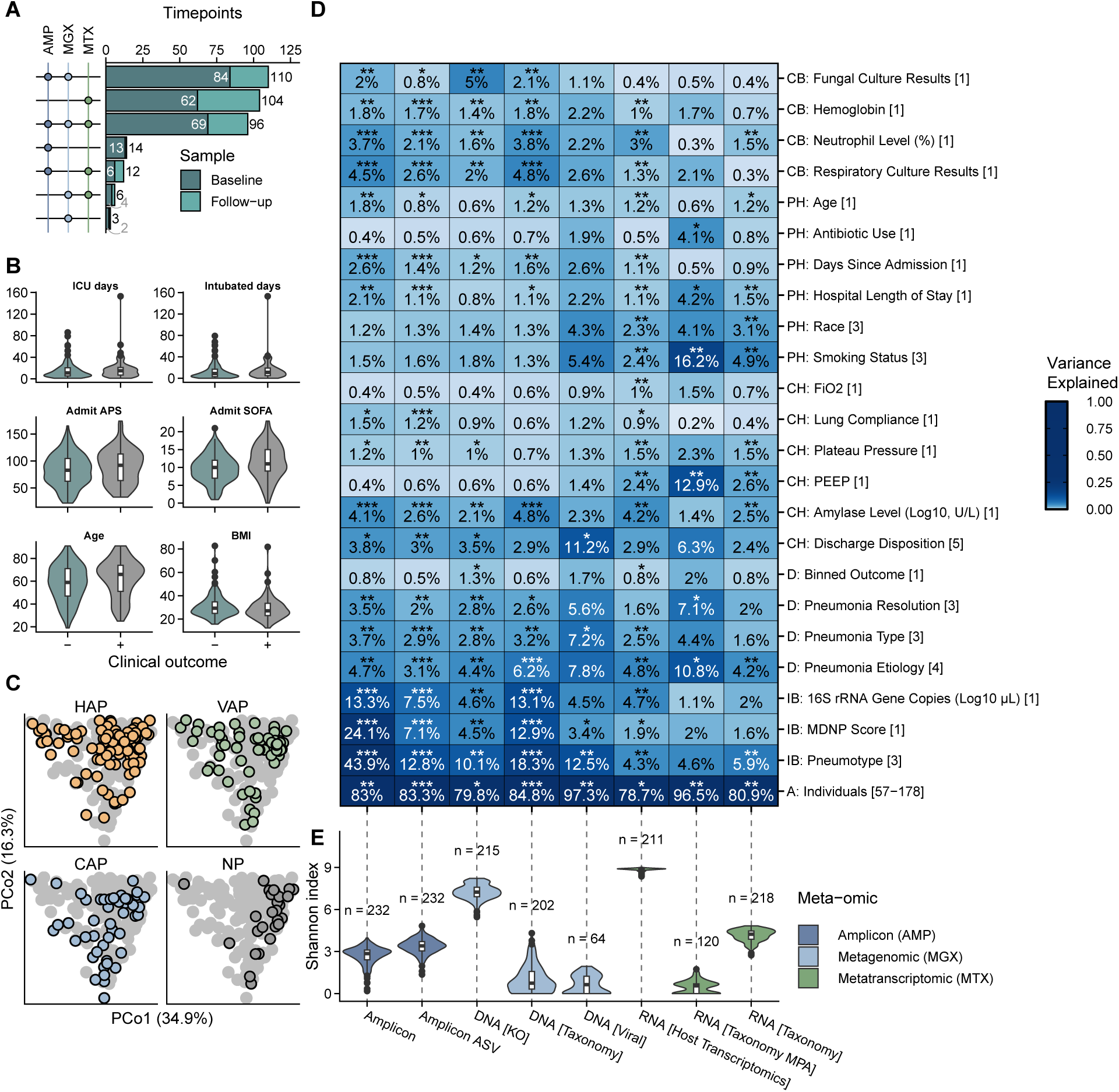
Multiomics of the lung microbial ecosystem during pneumonia reveals diverse associations with clinical features. (A) UpSet plot of multiomics sampling at the same time-point. Colors distinguish sample as either a baseline or follow-up BAL. (AMP = 16S rRNA gene amplicon, MGX = metagenomic, MTX = metatranscriptomic [including host-transcriptomics]) (B) Demographics of the SCRIPT cohort. Selected metadata features to provide quantitative overview of patient demographics. (- = negative binned clinical outcome [e.g., patient expires], + = positive binned clinical outcome [e.g., patient discharged and sent home]) (C) Principle coordinate analysis of the weighted UniFrac distance metric derived from amplicon profiles (genus-level). Colors are indicative of pneumonia category. Gray dots in the background are the shadow of all the points as if they were shown in a single plot rather than in small multiples. Percentages on axes are the variance explained by the given principle coordinate. See Figure S3 for remaining profiles. (HAP = hospital acquired pneumonia, VAP = ventilator-associated pneumonia, CAP = community acquired pneumonia, NP = critically-ill non-pneumonia control.) (D) Permutational multivariate analysis of variance analysis (PERMANOVA) quantifies the amount of variance in distance space explained by a given metadata features (e.g., pneumonia category) and tests for significance association. Percentages and color represent variance explained (R^2^). Columns are the different multiomic profiles. Bracketed numbers on right of y-axis metadata labels represent degrees of freedom. Significant association with high variance explained indicates metadata features as drivers of variation in the gene-expression or microbiota landscape. Features were nominally grouped into 6 categories: cellular biomarkers (CB), patient hallmarks (PH), clinical hallmarks (CH), disease (D), intrinsic biomarkers (IB), and an all (A) category for individuals. (* FDR *P* < 0.05, ** FDR *P* < 0.01, *** FDR *P* < 0.001; MDNP score = mean dissimilarity to non-pneumonia, PEEP = positive-end expiratory pressure, FiO2 = fraction of inspired oxygen, Binned Outcome = positive or negative discharge status as in (B)). (E) Shannon diversity of different multiomics profiles. 16S rRNA gene amplicon sequencing profiles include: Amplicon (genus-level) and Amplicon ASV (ASV-level); shotgun metagenomic profiles include: DNA [KO] (gene-content based on KEGG orthology terms), DNA [Taxonomy] (species-level), and DNA [Viral] (putative bacteriophages); and transcriptomic profiles include: RNA [Host Transcriptomics] (alveolar macrophage gene-transcript-expression), RNA [Taxonomy MPA] (species-level using MetaPhlAn4). (**Boxplot configuration**: Center line = median, box limits = upper and lower quartiles, whiskers = 1.5x interquartile range, points = outliers.)

The distribution of select clinical indicators of disease severity and risk were visualized to broadly capture the patient health profiles (Figure 1B). Clinical indicators include ICU days, intubated days, admit acute physiology score (APS), admit sequential organ failure assessment score (SOFA), age, and body mass index (BMI). These quantitative indicators are largely similar between patients independent of their binned, clinical outcome (Figure 1B). Note that binned outcome is based on discharge status and is distinct from pneumonia resolution (i.e., therapy success; see methods for detailed explanation).

### Drivers of gene expression and the microbiota landscape

To identify clinical features associated with variation in the microbiota and gene expression landscapes, permutational analysis of variance (PERMANOVA) analysis was performed comparing relevant distance space of omics features to clinical features and metadata suspected to be indicative of clinical outcome (Figure 1A). Features were nominally grouped into 6 categories: cellular biomarkers (CB), patient hallmarks (PH), clinical hallmarks (CH), disease (D), intrinsic biomarkers (IB), and an all (A) category for individuals. Explained variance is the square of the sum of squares statistic from PERMANOVA analysis. Order of features was determined by the rowise mean of the variance explained within each group. Overall, most significantly associated features (false discovery rate (FDR) *P* < 0.05) explain relatively little variation in distance-space (1-3%). Inter-individual variation explains the greatest amount of the variance in the data (Figure 1D, row “A: Individuals”), suggesting that personal molecular signatures are critical in disentangling pneumonia pathogenesis (Figure 1). Intrinsic biomarkers show the second greatest explained variance in the data with the greatest associations being detected between pneumotype and amplicon profiles (Figure 1D).

DNA-based approaches quantifying microbial features, either at the whole microbiome or bacteriome level, tend to similarly associate with clinical metadata features (Figure 1D), as expected by the covariation indicated by Mantel tests (Figure S2). Metatranscriptomic and host transcriptomic features do not consistently share the same feature similarity trends of DNA-based landscapes. Pneumonia category, which includes NP, CAP, VAP, and HAP, associates with amplicon (FDR *P* < 0.01) and shotgun taxonomic profiles (FDR *P* < 0.001) as well as shotgun functional profiles (FDR *P* < 0.01), indicating differences in microbial community structure and gene content landscape between patients with different pneumonia diagnoses (Figure 1C-D). A principle coordinate analysis visualizing these differences in amplicon data are highlighted in (Figure 1D). Pneumonia states can be further subcategorized by pathogen etiology: bacterial pneumonia, viral pneumonia, bacterial-viral pneumonia (i.e., superinfection), pneumonia of unknown etiology, or non-pneumonia. Pneumonia etiology associates with every tested profile with the exception of the putative virome, indicating a strong relationship to be explored regarding host-microbiome dynamics and clinical outcomes (Figure 1).

Shannon diversity index provide an overview of the evenness and richness of features between profiles. Gene-based profiles including KEGG orthology profiles from metagenomic sequencing and gene expression profiles from host transcriptomics are greater than organism-level profiles (Figure 1E). To assess the effect of processing pipeline in our analysis, we included amplicon profiles at the ASV-level and further glommed to the genus-level. Additionally we compared metatranscriptomic profiles derived from Bracken and MetaPhlAn4. Although we observe similar levels of shannon diversity between ASV-level and genus-level amplicon profiles, there is more pronounced variation between the two assayed metatranscriptomic profiles (Figure 1E).

### Quantifying microbiota landscape disruption during pneumonia

Changes to the microbiota landscape can be understood in at least two complementary ways: quantitative changes from a set baseline or control population (Figure 1C, Figure 2A-B) and identification of different microbiota states (Figure 3A-B).

**Figure 2.**
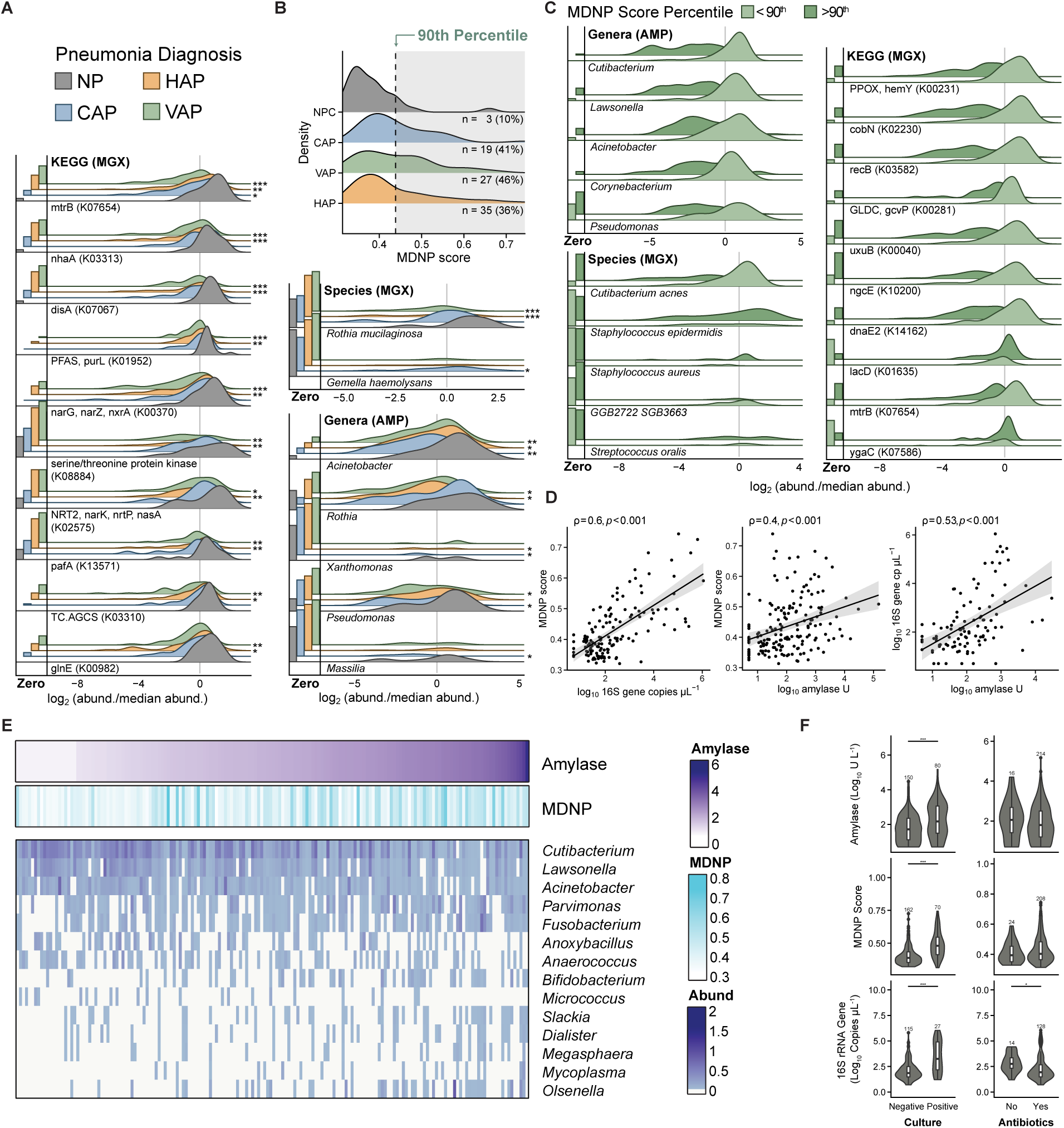
Pneumonia infection associates with an altered microbiota landscape indicative of aspiration-mediated disruption. (A) Abundance of the top differentially abundant (FDR *P* < 0.05) genes, species, and bacterial genera in each pneumonia category (i.e., HAP, VAP, CAP) relative to NP. Bar plots are the proportion of samples with zero-count therefore showcasing feature prevalence; bars are scaled such that touching the correspondingly colored line above indicates the feature was undetected in all samples for that group. Kernel distributions were calculated based on the subset of samples with detectable abundance after centering by the median and log_2_ transformation; heights are scaled by the proportion of detectable samples. Genes include their KEGG orthology term. (* FDR *P* < 0.05, ** FDR *P* < 0.01, *** FDR *P* < 0.001). (B) Distribution of the mean dissimilarity to non-pneumona (MDNP) score quantifying microbiome disruption relative to non-pneumonia control group. Score is calculated using the weighted UniFrac distance from amplicon profiles. The 90^th^ percentile of MDNP score within NP was used as a threshold to determine microbiota disruption in patients with pneumonia. (C) Abundance of the top differentially abundant (FDR *P* < 0.05) genes, species, and bacterial genera in disturbed microbial communities (>90^th^) relative to communities with structure typical of NP (<90^th^). Microbiome samples were binned into typical and disturbed subsets based on the 90^th^ percentile of MDNP score. Above this threshold, there is a 10% chance of a patient without pneumonia to have that particular arrangement of microbiota. (D) Relationship between bacterial load, amylase activity, and MDNP score. Shaded region represents 95% confidence interval. Statistics show Spearman rank correlation test. (E) Top differentially abundant genera (amplicon); samples ordered by increasing levels of amylase activity. (F) Distribution of bacterial load, amylase activity, and MDNP score binned by culture results and antibiotic usage at time of BAL. Stars represent statistical significance as determined by Wilcoxon test. (* FDR *P* < 0.05, ** FDR *P* < 0.01, *** FDR *P* < 0.001; **Boxplot configuration**: Center line = median, box limits = upper and lower quartiles, whiskers = 1.5x interquartile range, points = outliers.). **Acronyms**: HAP = hospital acquired pneumonia, VAP = ventilator-associated pneumonia, CAP = community acquired pneumonia, NP = critically-ill non-pneumonia control.

**Figure 3.**
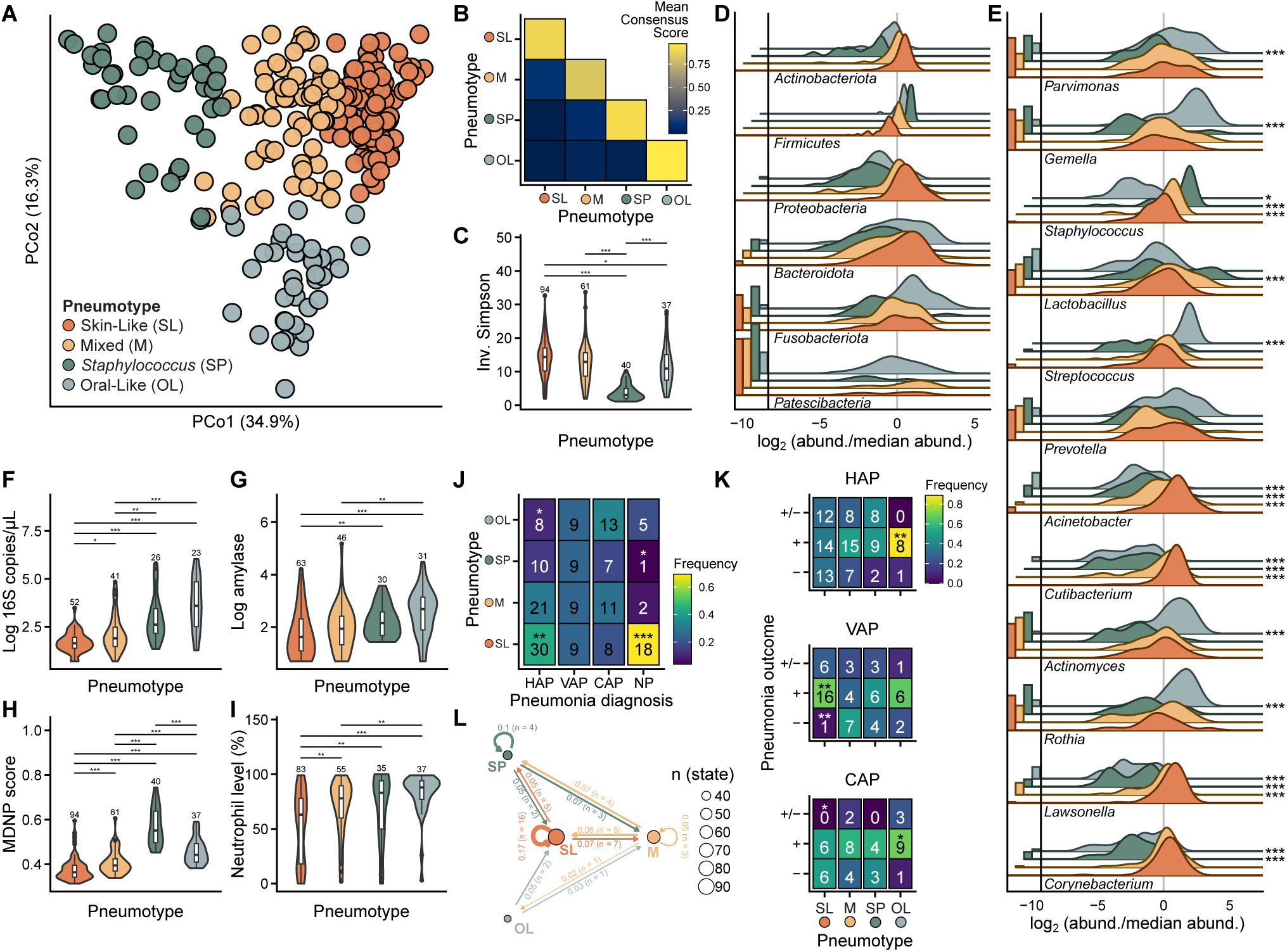
A posteriori identification of pneumotypes suggests stabilizing selective forces canalize community structure. (A) Ordination of weighted UniFrac distance derived from genus-level amplicon profiles. Colors represent the different microbiota states of the distal lung (i.e., pneumotypes) identified using cluster analysis. Percentages represent variance explained by the given principle coordinate axis. (B) Summary heatmap visualizing the mean cluster consensus score. Consensus clustering implementing the partition around medoids cluster algorithm was performed to determine number of groups evident in the weighted UniFrac distance space. (C) Trade-offs in diversity (Wilcoxon test) and (D) core phyla differentiate pneumotypes. (E) Most abundant taxa distinguish microbiota states. Taxa with a mean normalized abundance greater than 0.05 were selected (n=12). Stars represent the adjusted p-value of differential abundance analysis comparing pneumotype_M_, pneumotype_SP_, and pneumotype_OL_ to pneumotype_SL_. (F) Bacterial biomass, (G) amylase levels, (H) MDNP score, (I) and neutrophil abundance differ significantly between microbiota states. Pairwise comparisons show results of Wilcoxon test with Benjamini-Hochberg correction. (J) Heatmaps visualizing pneumotype frequency across pneumonia category (limited to baseline BAL) and (K) clinical outcome (includes baseline and follow-up BAL). Numbers in heatmaps are the count of BAL in each section; color of tiles is the proportion for that column. Stars represent the adjusted p-value of two-sided pairwise exact binomial tests used to determine deviations from expected distributions (i.e., evenly distributed across the column). Pneumonia therapy outcome is categorized as successful (+), indeterminate (+/-), and unsuccessful (-) treatment. (L) Frequency of transitions between pneumotypes. Nodes (circles) represent the different pneumotypes and the circle size is scaled to the number of samples. Edges (arrows) represent transitions between pneumotypes. Edge labels are the frequency of transitions between pneumotypes accounting for transition to outcome (i.e., final BAL are counted as transitioning to clinical outcome rather than to any pneumotype). (* FDR *P* < 0.05, ** FDR *P* < 0.01, *** FDR *P* < 0.001; **Pneumonia diagnosis**: HAP = hospital acquired pneumonia, VAP = ventilator-associated pneumonia, CAP = community acquired pneumonia, NP = critically-ill non-pneumonia control; **Pneumotypes**: SL = skin-like, M = mixed, SP = *Staphylococcus* predominant, OL = oral-like; **Pneumonia outcome**: - = unsuccessful treatment, + = successful treatment, +/- = indeterminate treatment; **Boxplot configuration**: Center line = median, box limits = upper and lower quartiles, whiskers = 1.5x interquartile range, points = outliers.)

#### Features distinguishing pneumonia from non-pneumonia

Differential abundance analysis was performed comparing the different pneumonia categories to non-pneumonia microbiota (Figure 2A). We report 100 genes (DNA [KO]), 6 genera (amplicon), and 2 species (DNA [Taxonomy]) as differentially abundant. Amplicon profiles indicate *Acinetobacter* is lower in all pneumonia categories (FDR *P* < 0.05 in HAP; FDR *P* < 0.01 in CAP, VAP). We observe lower levels of the oral-associated *Rothia* in VAP and HAP but not CAP relative to non-pneumonia (FDR *P* < 0.05); moreover *Rothia mucilaginosa* relative depletion associates with HAP and VAP microbiota profiles from shotgun metagenomics (FDR *P* < 0.001) but not in CAP profiles. *Gemella haemolysans*, another oral-associated microbe, is higher in CAP than in non-pneumonia (FDR *P* < 0.05). At the gene level, depletion of *mtrB*, a gene encoding a two component system response regulator protein involved in osmoprotection and cell proliferation, is associated with each of the pneumonia categories (Figure 2A). The *narK* and *narG* genes involved in nitrogen metabolism are relatively depleted in HAP and VAP (range FDR *P* < 0.05 - 0.001); relative depletion of the diadenylate cyclase gene *disA* is similarly depleted in these two categories (FDR *P* < 0.001).

#### Quantitative change from control population

Quantitative assessment of the microbial landscape gives relative directionality to a complex system dominated by individual signatures. Using 16S rRNA gene sequencing, we implemented this approach using BAL samples from critically-ill mechanically-ventilated patients who were adjudicated to be without pneumonia as a critically-ill population control (Figure 2B). Briefly, the mean dissimilarity to non-pneumonia (MDNP) score was determined for each sample by calculating the mean distance (i.e., weighted UniFrac) between a given sample and all NP samples. The 90^th^ percentile of MDNP score within the control group was used as a threshold to identify microbial profiles atypical in patients without pneumonia (Figure 2B, shaded region). For all pneumonia categories, 36-46% of samples were above the MDNP score 90^th^ percentile threshold. PERMANOVA analysis indicates that MDNP score strongly associates with 16S rRNA gene amplicon sequencing and shotgun metagenomic taxonomic and gene content profiles (Figure 1D). Below, we show the specific microbial hallmarks associated with elevated MDNP score.

#### Signatures of microbiome irregularity

To better understand the specific microbial features associated with extreme MDNP score (Figure 1D), we performed differential abundance testing as implemented in Maaslin2 (Figure 2C). We report 929 genes (DNA [KO]), 41 genera (amplicon), and 6 species (DNA [Taxonomy]) as differentially abundant in highly disrupted microbial communities (i.e., MDNP score greater than the 90^th^ MDNP percentile of patients with NP). Using the results from differential abundance testing, we visualized the top most significant features (FDR *P* < 0.05) (Figure 2). In bacterial profiles (Figure 2), we identified a trend in which microbiome profiles in the 90^th^ percentile of MDNP score associate with a lower overall abundance of several genera such as *Cutibacterium*, *Corynebacterium*, *Lawsonella*, *Acinetobacter*, and *Pseudomonas*. From shotgun metagenomic profiles, we observe elevated abundance of *Streptococcus oralis*, *Staphylococcus epidermidis* and, to a lesser degree, of *Staphylococcus aureus*. Relative depletion of *Cutibacterium acnes* and the uncultured *Lawsonellaceae* member GGB2722 SGB3663 is associated with profiles above the 90^th^ percentile of MDNP score. At the gene level, porphyrin biosynthesis genes *hemY* and *cobN* depletion are associated with microbiome disruption. Furthermore, *lacD*, encoding an inhibitor of *Streptococcus spp.* quorum sensing effectors regulating virulence and biofilm formation, and *ygaC*, encoding an uncharacterized gene regulated by Fur (iron), genes are elevated in microbiome disruption.

Differential abundance analysis of features associated with increasing levels of amylase reveals associations with 83 genes (DNA [KO]), 16 genera (amplicon), and 1 species (DNA [Taxonomy]). Associated genera are highlighted in Figure 2e. The abundance of *Slackia, Megasphaera, Dialister, Mycoplasma, Olsenella, Parvimonas, Fusobacterium, Bifidobacterium* are positively associated with amylase activity (range FDR *P* < 0.05 - 0.001). Furthermore, *Cutibacterium, Lawsonella, Acinetobacter, Escherichia-Shigella, Anoxybacillus, Anaerococcus, Micrococcus, Neisseriaceae* abundance negatively associates with amylase activity (Figure 2E).

### MDNP score is linked to elevated bacterial load and clinical markers of aspiration

Absolute bacterial load was measured using qPCR with a standard curve of known 16S rRNA gene sequence copy number. Amylase, an enzyme that constitutes up to 30% of salivary protein content, is a known marker for oral aspiration when detected in BAL fluid and a risk factor for pneumonia^13^. To test the hypothesis that aspiration events contribute to pneumonia pathogenesis by transmission of oral microbiota, we performed association testing between MDNP score, amylase activity, and bacterial load (Figure 2D). Using spearman rank order correlation, we identified monotonic relationships between MDNP score and 16S rRNA gene copy per *µ*L (*ρ* = 0.6, p < 0.001), MDNP score and amylase activity (*ρ* = 0.4, p < 0.001), and 16S rRNA gene copy per *µ*L and amylase activity (*ρ* = 0.53, p < 0.001). Based on these results shown in Figure 2B-D, we propose that MDNP score is a multivariate composite of pneumonia diagnosis and associated clinical features. Further analysis indicates that each of these hallmarks are elevated when BAL respiratory culture results are positive (Wilcoxon rank-sum test, p < 0.001; Figure 2F). The use of antibiotics is associated with lower bacterial load but not with amylase activity or MDNP score, although this analysis is underpowered as most patients were receiving antibiotics Figure 2F). These data are consistent with the hypothesis that microaspiration mediates changes in the lower respiratory tract microbiome. In addition, they suggest that pneumonia is associated with an increased overall bacterial load in the lungs.

#### Lung microbiota of critically ill patients exist in distinct pneumotype states

To test the hypothesis that conserved microbial communities comprise the lung microbiome during infection, we implemented an unsupervised machine learning approach (Figure 3). Clustering using partitioning around mediods incorporated phylogenetic similarity via the UniFrac distance; the number of clusters was determined using a consensus clustering approach (see methods for details) (Figure 3B). This approach identified four clusters of microbial communities, which are visualized in Figure 3A. The mean consensus score is visualized in Figure 3B. In total 261 samples were clustered into pneumotypes with varying microbial feature composition: Skin-like (pneumotype_SL_, 108 samples), mixed (pneumotype_M_, 70 samples), *Staphylococcus*-predominant (pneumotype_SP_, 40 samples), and oral-like (pneumotype_OL_, 43 samples).

### Microbial characteristics of pneumotypes

Alpha diversity quantified using the inverse Simpson index differs between the four pneumotypes Figure 3C. We report significant differences in diversity between pneumotype_SL_, pneumotype_M_, and pneumotype_OL_ to the singularly dominated pneumotype_SP_ using the Wilcoxon rank-sum test (FDR *P* < 0.001). Furthermore, pneumotype_SL_ displays somewhat greater diversity to pneumotype_OL_ (FDR *P* < 0.05).

To assess the distinguishing taxa between different pneumotypes, we conducted a differential abundance analysis at the genus and phylum level using Maaslin2^17^. Differential abundance tests were performed relative to pneumotype_SL_. Differential analysis reveals associations with 1743 genes (DNA [KO]), 63 genera (amplicon), and 14 species (DNA [Taxonomy]), and 9 taxa (amplicon) (Figure 3E).

Our results demonstrate significant tradeoffs in the relative abundance of phyla Actinobacteriota and Proteobacteriota with phyla Firmicutes and Fusobacteriota (Figure 3D). The phyla Actinobacteriota and Proteobacteria are significantly depleted in pneumotype_SP_, pneumotype_M_, pneumotype_OL_ while phylum Firmicutes is enriched (FDR *P* < 0.001). Additional tradeoffs in phyla abundance are also observed to a lesser degree. Bacteroidota is depleted in pneumotype_SP_ (FDR *P* < 0.001) and pneumotype_M_ (FDR *P* < 0.05). Pneumotype_OL_ is significantly enriched in phylum Fusobacteriota (FDR *P* < 0.001).

We identified two pneumotypes with a balanced yet distinguishable abundance of *Firmicutes* and *Actinobacteriota*, resembling pneumotypes previously observed in healthy volunteers (Figure 3D). One pneumotype exhibited enrichment of *Streptococcus*, *Gemella*, and other microbiota typically associated with the upper respiratory tract and oral niches (Figure 3E). This microbial profile corresponds to the “suppraglotic predominant”^9^ or “balanced”^8^ pneumotypes found in healthy lungs, which are associated with genera commonly involved in microaspiration events. We designate this pneumotype as pneumotype_OL_. Furthermore, pneumotype_SL_ is consistent with reports of “microbe depleted” or “background environmental predominant” states in healthy patients, resembling skin microbiota and exhibiting greater abundance of key markers such as *Corynebacteria*, *Cutibacteria*, and *Staphylococcus* than the other pneumotypes. Based on previous notions of contributions from the indoor environment and the prevalence presence of skin-associated microbiota on indoor surfaces^18^, we name this group pneumotype_SL_.

Pneumotypes dominated by a single phylum often associated with a single, predominant genus on a per-sample basis. Pneumotype_SP_ is primarily composed of genus *Staphylococcus* Figure 3E), with occasional contributions from other Firmicutes genera, such as *Lactobacillus* (FDR *P* < 0.001) and *Enterococcus* (FDR *P* < 0.05), in *Staphylococcus*-replete states (Figure S6). Pneumotype_SP_ likely overlaps with the previously identified pneumotype_SP_^8^, although, other genera contribute to the Firmicutes-dominated population structure. Pneumotype_M_ is predominately composed of *Staphylococcus*, *Corynebacteria*, and *Cutibacterium* which are genera commonly associated with the nares and skin niches^19,20^; additionally, this pneumotype is moderately abundant with microbiota associated with the human oral microbiome including *Streptococcus* (Figure 3). Although *Cutibacterium* is a prevalent contributor to pneumotype_M_ (prevalence = 61 samples), the genus is depleted relative to pneumotype_SL_ (FDR *P* < 0.001). The depletion of *Cutibacterium*, *Lawsonella* (FDR *P* < 0.001) (FDR *P* < 0.001), and *Acinetobacter* (FDR *P* < 0.001) along with the enrichment of *Staphylococcus* (FDR *P* < 0.001), *Granulicatella* (FDR *P* < 0.001), and maintenance of other oral microbiota is the distinguishing factor between between pneumotype_M_ and pneumotype_SL_ (Figure 3E).

### Pneumotypes capture aspiration-mediated neutrophil activation

Pneumotype association patterns indicate alternative mechanisms precede microbiome disruption and innate immune activation (Figure 3F-I). Pneumotype_OL_, followed by pneumotype_SP_, exhibits the highest bacterial load (Figure 3F), amylase activity (Figure 3G), and neutrophil levels (Figure 3I) among pneumtypes. MDNP score is overall greatest in pneumotype_SP_ while pneumotype_OL_ and pneumotype_M_ follow in descending order (Figure 3H). Therefore, pneumotype_SL_ displays low levels of bacterial load, amylase activity, microbiome disruption, and neutrophil activation. Furthermore, elevated neutrophil activation is present in pneumotype_M_ despite relatively low levels of microbiome disruption and putative aspiration. Thus, the pneumotypes capture varying levels of microbiome disruption associated alternating levels of aspiration and neutrophil activation.

#### Pneumotypes are enriched in a disease- and outcome-specific manner

To test the hypothesis that pneumotypes are distributed in a pneumonia category dependent manner at time of diagnosis, we implemented overrepresentation analysis using the pairwise binomial exact test compared to a null distribution (Figure 3J). Pneumotype_SL_ is enriched in HAP (FDR *P* < 0.01) and NP (FDR *P* < 0.001) populations. Pneumotype_OL_ is depleted in HAP (FDR *P* < 0.05) while pneumotype_SP_ is depleted in NP (FDR *P* < 0.05). VAP and CAP are not enriched or depleted for any particular pneumotype although CAP and NP appear to have a slightly higher, non-significant increase in pneumotype_OL_ compared to other pneumonia categories.

Furthermore, we tested if the distribution of pneumonia therapy outcome (i.e., successful, unsuccessful, and indeterminate treatment response) is associated with a specific pneumotype throughout treatment (Figure 3K). We report that although pneumotype_OL_ is rare in HAP (Figure 3J), it is associated with positive pneumonia therapy outcome (Figure 3K). Pneumotype_OL_ is also associated with successful pneumonia therapy in CAP. Despite an even distribution of pneumotypes in baseline VAP (Figure 3J), pneumotype_SL_ is indicative of positive clinical outcome (Figure 3K). Pneumotype_SL_ is also depleted in cases of indeterminate outcome in CAP (Figure 3K). Thus, pneumotype distribution at time of diagnosis is sometimes associated with pneumonia category and is indicative of therapy outcome in a context-specific manner.

#### Multiomic integration reveals complexity in the lung microbial ecosystem

Multi-omic network analysis provides insight into the lung microbial ecosystem (Figure 4). Inter-omic interactions were determined using Hierarchical All-against-All (HAllA) pattern discovery and subsequently visualized as a network (see methods for details). Hubs of highly connected nodes were identified based on the number of degrees; this led to the selection of eight nodes with a degree greater than 10. Hubs comprise the following amplicon features: *Streptococcus*, *Lawsonella*, *Staphylococcus*, *Rothia*, *Mogibacterium*, *Atopobium*, *Cutibacterium*; the following taxonomic shotgun features: *Streptococcus parasanguinis*, *Streptococcus mitis*, *Staphylococcus epidermidis*, *Streptococcus salivarius*, *Staphylococcus aureus*, *Gemella haemolysans*, *Streptococcus oralis*, *Corynebacterium striatum*, *Streptococcus anginosus*, *Lancefieldella parvula*, *Granulicatella* (SGB8255), *Streptococcus gordonii*, *Parvimonas micra*, *Cutibacterium acnes*; the following gene-level features: *ciaR* (K14983), *prdA* (K10793), *comE* (K12295), *ATPVG*, *ahaH*, *atpH* (K02107), *comX1/2* (K12296); and no features from other omic types. The network clearly clusters into three main groups with peripheral limbs (e.g., *Streptococcus/Rothia mucilaginosa* hub with *Atopobium* limb) and three additional singleton groups.

**Figure 4.**
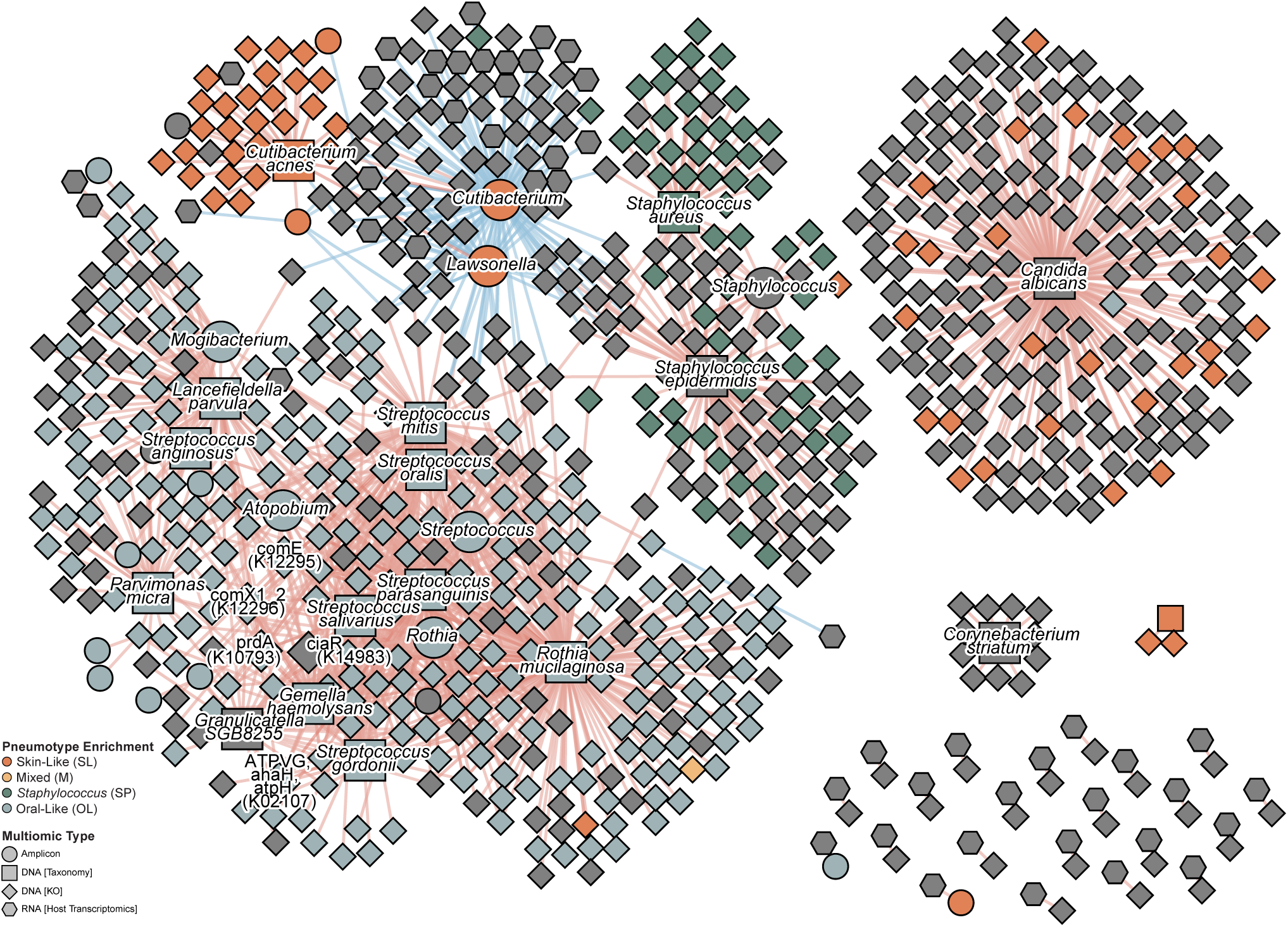
The lung microbial ecosystem is complex and rich with interactions across ecosystem components. Network visualization of associated omics features identified from HAllA. Edges are associations colored by Spearman rank correlation (red for positive and blue for negative); nodes are data features (e.g., genera, genes). Prevalent positive associations are observed between *Streptococcus* species and other oral microbiota (*Rothia spp., Gemella spp.*). Other major hubs include *Staphylococcus* and *Cutibacterium*. *Cutibacterium* negatively associates with many features including the expression of several host genes. Multiomics data integration includes feature profiles from four data types: shotgun metagenomic (taxonomic, functional KO profiles), 16S rRNA gene sequencing, and macrophage-sorted bulk RNA-sequencing (host transcriptomics, metatranscriptomic). Top significant associations from each dataset comparison are visualized (FDR *P* < 0.05). Nodes with at least 10 significant associations are highlighted in the network as hubs with slightly larger sizes. Nodes high in one pneumotype are colored accordingly (see methods for details). Features that were high in multiple groups or no groups were kept as gray. See Figure S7 for a fully labeled network diagram.

We observe co-abundance of oral microbiota, including many species of *Streptococcus* (*S. mitis*, *S., anginosus*, *S. oralis*, *S. gordonii*) in association with other oral microbiota (*Rothia spp.*, *Gemella spp.*). These interaction hubs are particularly evident in the central taxonomic clusters in the network visualization (Figure 4). *Cutibacterium*, a member previously identified in the environmental-like pneumotype of healthy individuals, is positively entangled with *Corynebacterium*, *Lawsonella*, and *Acinetobacter*. This group is typically negatively associated with features (e.g., KOs) that are positively associated other oral microbiota such as *Rothia* and *Streptococcus* species. Microbial markers of the skin-like microbiota state (e.g., *Cutibacterium*) negatively associate with expression of genes involved in inflammatory response (e.g., interleukin-1 beta), suggesting the skin-like state’s role as a baseline in the microbiota landscape (Figure S7).

*Staphylococcus* represents a third unique hub, typically negatively associating with the oral and *Cutibacterium* clusters described above (Figure 4). Amplicon analysis identifies positive correlation between *Staphylococcus* in amplicon and *Staphylococcus aureus* identified in shotgun metagenomics. *Staphylococcus epidermidis* is also present, but no connections are observed between it and the amplicon *Staphylococcus* node in the subset of top connections. This disconnect suggests that species- or strain-level differences in microbiome composition may have important implications for disease state or outcome.

## DISCUSSION

By profiling lung microbial ecosystem components through our different multiomics methods, we show that microbial community structure and host gene expression change in unison with clinical and physiological hallmarks of patient health. That is, we see variation in clinical phenotypes reflected in different component landscapes (Figure 1D). For example, pneumonia etiology associates with host gene expression and microbial community structure (Figure 1D, PCoA visualizations in Figure S4A-C,F). This cohesive association between system components suggests a concomitant response to a single stimulus. However, sometimes change in clinical phenotype is uniquely reflected in one system component. In this context, it suggests that system components may also function independently. There are likely mechanistic connections between lung microbiota and clinical phenotype that do not involve changes to host gene expression and vice-versa. While component landscapes are not always the same, we propose there are convergent regions that exhibit similar topography. We further hypothesize that the lung microbiome interacts with host gene expression and patient physiological status in a way that is relevant to pneumonia therapy outcome. The underlying mechanism and direction of causality (e.g., which component changes first) remain to be explored, but we can leverage these associations to build ecological models of pneumonia outcomes.

We bolster challenges presented against the classical clinical conceptualizations of pneumonia, which posit that a single pathogen comes to dominate the lung microbial ecosystem, by showcasing associations between microbiota state and infection-specific dysbiosis with physiological determinants^1,5,15^. “Individual” accounts for the greatest amount of variation (Figure 1D), yet conserved microbiota states called pneumotypes are indicative of pneumonia category and clinical outcome (Figure 3J-K). Pneumotype_SL_ (abundant with microbiota associated with nares and skin) is enriched in patients with HAP or without infection (NP) (Figure 3E,J). Neutrophilic activation is more varied with overall greater spread in pneumotype_SL_ than in the other pneumotypes (Figure 3I). The microbial composition of pneumotype_SL_ suggests a role for microbiota colonization from the hospital environment or skin microenvironment in patients with HAP or NP. Pneumotype_SL_-associated features negatively correlate with expression of genes typically involved in inflammatory response (Figure 4, Figure S7). Acquisition of low-abundant nosocomial pathogens without microbiota state disruption may distinguish pneumotype_SL_ in patients with HAP and NP. Pathogen presence would thus likely contribute to pneumonia symptom induction despite non-dominant levels.

Pneumotypes are predictive of therapeutic success in a category-dependent manner (Figure 3K). Although rare in patients with HAP, occupancy of an oral-like microbiota state (pneumotype_OL_) is indicative of successful pneumonia therapy in patients with HAP and CAP (Figure 3J,K). Pneumotype_OL_ associates with elevated amylase levels, local neutrophilic activation, and elevated bacterial load (Figure 3F,G,I). Amylase is typically only expressed by the pancreas and salivary glands. When abundantly detected in BAL, it is hypothesized to be indicative of aspiration from the oral cavity^13^. Indeed, increasing amylase levels associate with increasing oral-associated microbiota (Figure 2E). This association is especially noteworthy as many of the associated genera are anaerobic and often fastidious (e.g., *Fusobacteria*, *Parvimonas*, *Slackia*). This association would likely be undetectable in typical culture-based measurements. We implement MDNP score, a measurement of infection-specific dysbiosis based on previous implementations of dysbiosis quantification in the gut (Figure 2B)^21^. *Staphylococcus aureus*, *Staphylococcus epidermidis*, and *Streptococcus oralis* are relatively enriched in highly disturbed microbial communities determined via MDNP score. We report co-associations between MDNP score, amylase levels, and bacterial load (Figure 2D). Together, these data suggest aspiration-driven disruption as an underlying mechanism of pneumotype_OL_ assembly.

The relationship between lower respiratory tract microbiota and other human and environmental microbial niches remains an open field of investigation. Lung community structure is thought to be a balance between microbial immigration (e.g., from the air and oral cavity) and microbial elimination (e.g., mucociliary clearence, immune landscape) where few microbes actually take up residence^1^. During health, this relationship is thought to lead to neutral (stochastic) assembly of a low-biomass microbiome^22,23^. The lungs are a meeting ground for many microbial community inputs^24,25^. Microbial elimination forces are thought to confer resilience to community coalescence. However, disruption of the underlying ecological forces likely contributes to changes in how the community assembles. Deterministic assembly may therefore arise in the lung should environmental conditions begin to favor regional microbial growth, either because a new microbe takes advantage of a particular niche or the lung environment itself changes. Mechanistic connection between the oral microbiome is of particular interest due to the observation of oral microbiota in the lower respiratory tract in health^9^. Here we report a dynamic relationship between microbial landscape disruption (MDNP score) and suspected aspiration (high BAL fluid amylase levels) during pneumonia (Figure 2D). These data support the hypothesis that oral aspiration events contribute to lower respiratory tract bacterial load and promote transitions to disturbed microbiota states. We hypothesize that this relationship exists as a function dependent on the number or volume of aspiration events. Therefore, frequent or large aspirations events may yield altered microbiome. Oral microbiota are known to activate innate immunity in the lower respiratory tract^9,11,12^. Low-level exposure during health may prime the immune system for better anamnestic response during pneumonia-inducing exposures, explaining pneumotype_OL_’s association with pneumonia resolution.

We further explored this relationship by examining microbiota state stability throughout hospitalization in patients with serial longitudinal samples. Patients with pneumotype_SL_ stay in the same state more often than patients with different microbiota states (Figure 3L). This suggests pneumotype_SL_ may be more stable than other states. In contrast, pneumotype_OL_ is more often the first and final BAL before (positive) outcome determination than other pneumotypes (Figure 3K,L); patients with this pneumotype do not often have serial samples making measurable transitions to other pneumotypes or maintenance rare. Patients with pneumotype_SP_ and pneumotype_M_ exhibit an intermediate amount of state transition and retention. There are no detected instances of direct transition to pneumotype_OL_ from pneumotype_SP_ (Figure 3L). This may be due to antagonistic relationships between members of each pneumotype, as has been mechanistically explored between *Staphylococcus aureus* and *Fusobacterium nucleatum* in the nares^26–28^.

In addition to specific taxa, several genes encoding nitrogen metabolizing functions are relatively depleted in metagenomes from patients with HAP or VAP (Figure 2A). Activated macrophages, neutrophils, and other inflammatory cells produce reactive nitrogen species as inflammatory byproducts^29–33^. It has been proposed that respiratory tract microbiota, notably *Pseudomonas* and other Gammaproteobacteria, may use these nitrogen species as terminal electron acceptors^34,35^. In the gut, the use of these inflammatory byproducts by commensals has been empirically demonstrated to confer a competitive advantage^36^. In this study, *Pseudomonas* and *Acinetobacter* follow a similar, albeit not perfectly consistent, trend to nitrogen metabolizing genes; these genera are relatively depleted in various pneumonia categories compared to NP (Figure 2A). Additionally, the pneumotype_OL_ member *Rothia mucilaginosa* strongly follows this trend and is also known to encode nitrogen metabolizing genes (Figure 2A)^37,38^. Indeed, *R. mucilaginosa* is significantly associated (FDR *P* < 0.05) with abundance of the *nhaA* (K03313) and *narK* (K02575) genes involved in nitrogen metabolism (Figure S7). This indicates *R. mucilaginosa* at least partially drives nitrogen metabolism gene abundance in our data. Our results are consistent with the idea that nitrogen metabolism may confer some selective advantage to *R. mucilaginosa* and other nitrogen-competent bacteria in the lung environment of critically ill patients without (nosocomial) pneumonia. Specifically, pneumotype_OL_’s characteristically high neutrophil load may support *R. mucilaginosa* growth through the release of reactive nitrogen species during innate response (Figure 3I). This effect may be present in patients with nosocomial pneumonia but overshadowed by stronger selective forces, e.g., antimicrobial therapy. Future research implementing metabolomic studies and nitrogen flux analysis may shed further light on this hypothesized phenomenon.

We hypothesize that microbiota state transition is an underlying mechanism of successful response to pneumonia therapy. Moreover, completely inert states may challenge therapy success. This may be due to greater stability or resilience of the pathologic microbial community to (insufficient) selective forces conferred from treatment. Additionally, interpretation of clinical data may be more challenging with unyielding microbial communities as microbiologic testing may be insufficient. An important limitation of our study is that longitudinal analysis of BAL specimens from patients in the ICU suffers from sampling bias, as typically the sickest patients expire and healthiest patients recover prior to repeat samples, excluding them from representation. Therefore, our longitudinal samples split by pneumonia resolution and failure to respond to therapy likely exclude the extremes of response, resulting in potentially greater overlap.

Based on our snapshot and longitudinal analysis findings, we hypothesize that the lower respiratory tract microbiome proceeds through distinct pathways during pneumonia progression and resolution. Together these data are consistent with two models of pneumonia onset and treatment progression (illustrative summary in Figure S11). First is a regressive-state model. Pneumonia onset may occur such that the global microbiota state remains similar to that of patients without infection. Therapeutic intervention decreases infection-specific signatures despite an inert microbiota state. Second is a non-regressive-state model. Probable disruption events prior to diagnosis drive a highly perturbed microbiota state. Therapeutic intervention alters the landscape; the destabilized microbiota state progresses towards a new minimum. Existing experiments show that pulmonary inoculation with a mock oral community (low-complexity) produces transient effects on community structure in a murine model^39^. The effects of treatment and community complexity remain underexplored. Future research aimed at empirically testing these proposed dynamics *in vitro* and *in vivo* is necessary.

As research focusing on large-scale center-wide studies emerges, our understanding of the temporal dynamics of the lung microbiome will continue to expand. This work will help redefine our understanding of pneumonia, further allowing the classification of heterogeneous etiologies and disease substates. Eventually, information about the lung microbiome will enable finer diagnostics and mid-treatment evaluation of prognosis, eventually leading to radically improved pneumonia therapies.

## METHODS

### Sample acquisition and clinical adjudication

Samples from patients were collected from participants enrolled in Successful Clinical Response In Pneumonia Therapy (SCRIPT) study STU00204868 which obtained approval from the Northwestern University Institutional Review Board. Sampling of the lower respiratory tract via nonbronchoscopic and bronchoscopic bronchoalveolar lavage (NBBAL and BAL) is routinely performed in mechanically ventilated patients in the intensive care unit (ICU) at our institution. Per our BAL protocol, clinicians use a disposable bronchoscope to inspect the airway and wedge the scope in the airway segment that corresponds to a radiographic infiltrate or where secretions suggestive of pneumonia are present. After the bronchoscope is wedged, 120 cc of saline is instilled through the scope in four aliquots. After discarding return on the first aliquot, subsequent return volume is sent for clinical studies including semi-quantitative bacterial culture, multiplex PCR, cell count and differential. Frequently, fungal studies and amylase levels are also obtained by the clinical team. Participants enrolled in the SCRIPT study had residual BAL fluid retrieved and multicolor flow cytometry performed within 24 hours of the procedure; various samples were then aliquoted and stored frozen at -80*^◦^*C in 1 mL aliquots for later processing. In addition, the hospital courses of all patients enrolled in SCRIPT are adjudicated by a panel of six pulmonary and critical care physicians to achieve consensus on the diagnosis of pneumonia, the clinical state of the patient at various time points during treatment of the pneumonia episode, and the overall outcome of the patient’s hospitalization. The adjudication protocol and results have been published^16^. Relevant to this study, an overall outcome of ‘success’ is designated to patients who survived the duration of treatment and experienced improvement in ventilator requirements and markers of infection. An overall outcome of ‘failure’ is given to patients who continued to require antibiotics, had evidence of persistent infection/inflammation, or did not survive the completion of pneumonia treatment. Aliquots which were successfully processed for sequencing but for which patient metadata could not be mapped with certainty (n=32 BAL) or the patient(s) later withdrew from the study (n=2 BAL) were excluded from analysis and visualizations. In cases where the BAL were from lung transplant recipients (n=3 patients), metadata were often limited requiring exclusion in most analyses. At the end of the entire processing pipeline, we yielded clean data for 232 amplicon, 202 metagenomic [Taxonomy], 215 metagenomic [KEGG Orthology], 64 metagenomic [Viral], 218 metatranscriptomic [Taxonomy from Kraken/Bracken], 119 metatranscriptomic [Taxonomy from MetaPhlAn], and 210 transcriptomic [Host Transcriptomics] profiles derived from 345 BAL samples.

### Metagenomic DNA extraction

Frozen sample aliquots were thawed and processed using the MolYsis Complete 5 kit (Order No. D321-050, D-321-100) for DNA purification and host depletion. Briefly, host cells are disrupted using chaotropic salts and extracellular DNA is digested using the MolB DNase enzyme, which is robust against inhibitors. DNase is inactivated and microbial cells are lysed for spin-column-based DNA purification. DNA concentration was assessed using a Qubit fluorometer (Invitrogen). Metagenomic DNA size was quality controlled using a TapeStation genomic DNA assay.

### Shotgun metagenomic library construction and sequencing

Shotgun metagenomic libraries were prepared using NEBNext® Ultra™ II FS DNA Library Prep Kit for Illumina (NEB Catalog E7805L) following manufacturers’ instructions. Library quality and quantity are measured respectively by Tapestation (HSD1000 Agilent Technologies) and Qubit fluorometer (Invitrogen). Libraries were pooled in an equimolar ratio for multiplexed sequencing. Samples were omitted from pooling in cases where libraries were not detected. Pooled libraries were submitted for sequencing at the University of Illinois-Chicago Genome Research Division Sequencing Core. Sequencing was performed on a NovaSeq instrument usng 2x150bp paired-end chemistry.

### Shotgun metagenomic data processing

Shotgun metagenomic sequencing data were adapter and quality trimmed using fastp (v.0.23)^40^. Low complexity sequences were filtered using bbduk (entropy threshold = 0.3) from the BBMap software suite (v.39.01) to filter reads likely originating from human genomic DNA missed during *in silico* removal (i.e. alignment)^41^. High quality, complexity-filtered reads were then aligned to the human reference genome (CHM13 Telomere-to-Telomere with Y chromosome from GRCh38) using bowtie2 (v.2.4.5) with ‘—very-sensitive’ parameters^42^. Using samtools (v.1.10.1), unmapped read pairs (-f 12 -F 256) were selected for downstream analysis. Reads were processed using MetaPhlAn4 (v.4.1.0)^43^ to assess taxonomic composition (mpa_vJun23_CHOCOPhlAnSGB_202403 version database). Species-level MetaPhlAn4 profiles were filtered to only include features observed in at least 2 samples (n=224). Profiles were then normalized using total sum scaling followed by AST normalization. Functional metagenomic profiles were determined using HUMAnN3 (v.3.9) (–translated-subject-coverage-threshold 0.0 –nucleotide-subject-coverage-threshold 0.0 –bowtie-options=“–very-sensitive-local”)^43^. Reads were mapped to the ChocoPhlAn database (v.201901_v31) using nucleotide search; unmapped reads were then processed using the UniRef90 database with translated search. Gene family abundances, which are default in read-per-kilobase, were then normalized to counts-per-million. Normalized abundance profiles were then regrouped to KEGG orthology (KO) terms for downstream analysis. For KO profiles, unmapped and ungrouped categories were dropped prior to total sum scaling and AST normalization.

#### Viral analysis pipeline

Putative phage contigs were identified using geNomad (v.1.5.2) with default parameters^44^. Viral contigs were checked for completeness using CheckV (v.1.0.1)^45^. Contigs identified as viral by geNomad were aligned to each other using megablast. Alignments were used to cluster viral contigs at 95% nucleotide identity and 85% alignment fraction to create representative vOTUs. ANI calculation and clustering were done using anicalc.py and aniclust.py, respectively, from the CheckV GitHub repository. The longest sequence was selected from each cluster as the representative for each vOTU. The vOTUs that were designated as medium quality, high quality and complete by CheckV were kept for downstream analysis. To determine abundance of vOTUs across samples, cleaned reads from all samples were first aligned to representative vOTUs using BBMap (v.39.01) with the flag: -ambiguous=best^46^. Metapop (v.0.0.42) was used to create an abundance table^47^. Raw abundance was calculated as the average sequencing depth truncated to the central 80% (termed as TAD). Phage host predictions were made using iPHoP (v.1.3.2)^48^. The network created from iPhoP outputs mapped vOTUs to the most likely host based on multiple phage host pairing tools. Viral cluster network and phage host interaction network were visualized using Cytoscape (v.3.9.1) (Figure S9)^49^.

### 16S rRNA gene amplicon library construction and sequencing

To assess the composition of the lung microbiome, we conducted 16S rRNA gene amplicon sequencing on 261 bronchoalveolar lavage fluid (BALF) samples. A total of 6 water controls and 3 Zymo-BIOMICS Microbial Community DNA Standards (cat no. D6305) were included. Library preparation was performed using a semi-automated adaptation of Illumina’s recommended approach. Briefly, 18 µL per sample were aliquoted into a 96 well plate and vacuum centrifuged to a dry pellet. DNA pellets were resuspended with nuclease-free water to 1.25 ng/µL or to a maximum volume of 10 uL using a dragonfly liquid handler. Primary amplification of the V3/V4 rRNA gene regions was performed using universal primers, 341F and 805R, with Illumina adapter regions on the 5’ end (F-TCGTCGGCAGCGTCAGATGTGTATAAGAGACAGCCTACGGGNGGCWGCAG, R-GTCTCGTGGGCTCGGAGA Primary amplification reactions were prepared using 10 µL of concentrated DNA or water for no template controls, 12.5 µL of 2x KAPA HiFi HotStart Ready Mix (cat no. KK2602), and 2.5 µL of primer mix (2 µM of forward and reverse primer). Secondary amplification to attach Illumina indexes was performed using IDT for Illumina DNA/RNA UD Indexes kit with the same KAPA HiFi HotStart polymerase. SPRI bead cleanups were performed between each amplification step. Expected library size was assessed using the TapeStation High Sensitivity D1000 capillary fluorescence assay. Libraries prepared from water (negative) controls were still included in the sequencing pool despite undetectable TapeStation traces to ensure sequencing of low level background contaminants. Library were pooled and sequenced twice on an Illumina NextSeq 2000 instrument with the 2x300 bp P1 Reagents kit (cat no. 20075294).

### 16S rRNA gene amplicon sequencing data processing

#### ASV denoising and preliminary filtering

Amplicon data were demultiplexed using BCL convert (v.4.0.3); all samples and six out of seven no template controls were able to be demultiplexed. Next, reads were adapter-trimmed using fastp (v.0.23)^40^. Custom scripts using the QIIME2 platform (v.2021.11) were used for pipeline analysis^50^. Amplicon sequence variants (ASVs) were denoised using the DADA2 algorithm. A phylogenetic tree was constructed using “align-to-tree-mafft-fasttree” in QIIME2. ASVs were then taxonomically classified using the plugin “feature-classifier classify-consensus-vsearch” with the Silva 138 SSURef NR99 full-length database as a reference^51,52^. Downstream analysis was performed using R (v.4.2.3) and RStudio (v.2023.6.0.421). QIIME2 objects were loaded into R as a phyloseq object with the qiime2R package (v.0.99.6). ASVs with a kingdom-level assignment of Eukaryota or Unassigned were removed; ASVs with a genus-level assignment of Chloroplast or Mitochondria were also removed. This filtering yielded 11,344 ASVs from the original count of 22,275 ASV. This filtering represents approximately half of the denoised ASVs but only a negligible amount of the total reads. Then 46 additional ASVs with no counts were removed. Putative contaminating ASVs were identified from the six demultiplexed no-template controls using the Decontam package (v.1.18.0)^53^. The prevalence method with a probability threshold of 0.05 was used in Decontam. Of the 11,298 ASVs, 48 were identified as putative contaminants. Additional ASVs only found in control samples (i.e., water controls and Zymo standards) were then filtered out for downstream analysis of BAL samples (n=47). Final read count for cleaned sample data ranged from 39,922 to 664,319 reads.

#### Data normalization

ASV-level normalization and genus-level normalization were performed independently. For ASV-level normalization, ASVs were first filtered by a minimum read count of 2 in at least 5 samples, leaving 957 ASVs. Abundance was then normalized using total sum scaling followed by arcsine square-root transformation (AST) for variance stabilization. At the genus level, taxa names were merged using the tax_glom function in phyloseq; taxa without an assigned name at the genus level were dropped (default parameter NArm=TRUE). After this step, 710 genera were present. Low-abundant genera were filtered using a minimum read count of 2 in at least 2 samples, leading to a final count of 461 genera. Genus-level data were then normalized using total sum scaling and arcsine square-root transformation. A stricter prevalence filter was chosen for ASV-level filtering to balance data sparsity potentially derived from sequencing and pipeline noise, e.g., splitting of copy number variants from the same organism into multiple ASV groups.

#### Quantitative PCR

Quantitative PCR was performed with universal primers targeting the 16S rRNA gene to determine absolute bacterial load in BALF samples^54^. Reactions contained 10 µL 2x PowerUp SYBR Green Master Mix (Applied Biosystems, Cat no. A25741), 9 µL of nuclease-free water (Invitrogen, Cat no. AM9932), and 1 µL of DNA template with a final primer concentration of 400 µM forward and reverse primer. Thermocycling was performed using a QuantStudio3 under the following conditions: 50°C for 2 minutes, 95°C for 10 minutes, followed by 40 cycles of 95°C for 15 seconds and 60°C for 1 minute. A previously constructed plasmid containing a 167 bp target region was serially diluted to make a standard curve of known gene sequence copies (10^1^ to 10^7^)^55^. Samples with detectable signal below the limit of quantification were considered to be one-half the value of the limit of quantification (i.e., 5 copies/reaction). Up to nine no template controls were included per plate. Reaction plates and standard curves were prepared using an EpMotion5073M Liquid Handler (Eppendorf).

#### Transcriptome sequencing

Bulk RNA sequencing was performed on alveolar macrophages recovered from bronchoalveolar lavage sequencing using fluorescence-activated cell sorting as previously described^56^. Briefly, total RNA was extracted from samples followed by ribosomal RNA depletion. Sequencing libraries were prepared using using a reverse-stranded protocol and sequenced on a NextSeq2000 to produce 75 bp single-ended reads.

#### Transcriptome sequencing data processing

Gene expression tables were generated using a standard netflow workflow as previously described^56^. Fragments Per Kilobase of transcript per Million mapped reads (FPKM) were used for downstream analysis. The expression table was limited to protein coding genes; protein coding genes were first identified using the biomaRt package (v.2.54.1) and selecting genes for which the “gene_biotype” was encoded as “protein_coding”. Then, a prevalence filter was applied requiring gene RNA product expression detection in at least 20 samples. Expression tables were then re-normalized using total sum scaling followed by arcsine square-root transformation for variance stabilization. The Bray-Curtis distance was calculated using the vegdist function from the vegan package (v.2.6-4).

Unmapped reads were processed for taxonomic profiles. Reads were processed using MetaPhlAn4 (v.4.0.6, mpa_vOct22_CHOCOPhlAnSGB_202212 database). Profiles were assessed at the genus level and features detected in greater than one sample were retained (n=34). Unclassified reads features was removed prior to total sum scaling and AST normalization. Complementary to marker-based analysis, taxonomic profiling was additionally performed using Kraken2 (v.2.1.2) using the standard database followed by relative abundance estimation using Bracken (v.2.7.0; -t 10 -l ‘S’ -r 75)^57–59^. Features which were not detected at a threshold of 0.001 abundance in at least 10 samples were excluded (remaining n = 302 features) prior to AST normalization.

### Meta-omic data integration

We implemented a pairwise network structure using HAllA (v.0.8.20)^60^. Data types were subset to their shared number of samples and low prevalence features were excluded (<10%) prior to being processed using HAllA. Significant features were selected for using an alpha threshold of 0.05; associations were quantified using the Spearman coefficient. For constructing the network, interaction pairs were thresholded by association (Spearman’s rho > 0.5) and significance (FDR *P* < 0.05). Features meeting these criteria and occuring in a HAllA-identified cluster were selected for visualization, leading to 820 nodes (features) with 1398 edges (interaction). Nodes with greater than or equal to 10 degrees were highlighted in the network visualization as hubs with slightly larger sizes. Network was visualized using Cytoscape (v.3.10.1) using the edge-weighted spring embedded layout with the association strength as the weight. Overlaps were removed and nodes shape was by datatype (e.g., amplicon profile). Nodes were colored by features that were differentially over-abundant in pneumotypes; negative associations were considered to be “high” in pneumotype_SL_ as it was the baseline comparison group. Features that were high in multiple groups were kept as gray.

### Statistical analysis

#### PERMANOVA

For each metadata field, samples without recorded metadata were dropped. Samples that were present in the filtered metadata table and distance matrix were kept for PERMANOVA analysis. PERMANOVA analysis was performed using the adonis2 function in the R package vegan; a total of 4,499 permutations were performed for each test. Multiplicity correction was performed using the Benjamini-Hochberg method for each dataset.

#### Mantel test

To test for covariation between multiomics profiles, pairwise comparisons using the Mantel test were performed between each data set. Distance matrices were subset by the intersection of samples in each multiomics distance matrix. For instance, amplicon and metagenomics distance matrices were subset to include only the samples present in both matrices. The Mantel test was performed using the mantel.rtest function from the ade4 package. Multiplicity correction was performed using the Benjamini-Hochberg method.

#### Differential abundance testing

Differential abundance was tested using Maaslin2 (v.1.12.0)^17^. Features with a prevalence of less than 10% were removed before significance testing. Abundance profiles were AST normalized before evaluation.

#### MDNP analyses

Mean dissimilarity to non-pneumonia (MDNP) scores were determined using genus-level amplicon profiles. The mean Weighted UniFrac distance was calculated between each sample and the entire NP population. The 90^th^ percentile of MDNP score within NP was used to determine highly irregular microbial communities. At the 90^th^ percentile threshold, samples only have a 10% chance of having a similar arrangement of bacterial features to NP microbiome profiles.

#### ZLR plot visualizations

Zero log ridge plots were made to visualize differentially abundant microbial features. Bar plots on the left-hand side indicate detectable prevalence. Bar plots are scaled such that samples entirely undetected in a given category will reach the respective category baseline in the feature above it. Kernel density estimation plots were implemented using the ggridges package (v.0.5.6). Distributions were calculated using the ‘density_ridges’ implementation on data centered on (i.e., relative to) the median detectable abundance. Maximum height was scaled by the proportion of the total number of zero counts.

#### Cluster identification

To test the hypothesis that lung microbiota exhibit distinct pneumotype states, we developed an approach that incorporates phylogenetic relatedness and cluster stability. Integrating phylogenetic relatedness into cluster identification increases the likelihood of linking distinct population structures to shifts in ecological states or microenvironmental conditions, as closely related taxa have a greater tendency to fulfill similar niches^61^. Prior to cluster analysis, samples were normalized at the genus level using total sum scaling with arcsine square transformation. We used weighted UniFrac distance, a comprehensive measure that combines phylogenetic relatedness and relative abundance, to assess pairwise sample similarity^61^. Unsupervised learning was conducted through consensus clustering with iterative sample permutation, utilizing the weighted UniFrac metric to identify stable clusters as implemented in ConsensusClusterPlus (v.1.62.0)^62^. This methodology yielded four stable clusters representing putative pneumotypes.

#### Frequency tests

Unless otherwise indicated, violin plots with significance testing were visualized using the geom_pwc package from the ggpubr package (v.0.6.0). Pairwise Wilcoxon sign-rank test analyses were performed as implemented in the rstatix package (v.0.7.2) followed by Benjamini-Hochberg correction.

#### Overrepresentation analysis

Overrepresentation analysis was performed using a pairwise binomial distribution test against an expected probability. The test was performed as implemented in the rstatix package using the ‘pairwise_chisq_test_against_p’ function. The expected probability comparisons of the microbiome state distribution among pneumonia subtype was compared to the null distribution. As pneumonia therapy is hypothesized to affect microbiome composition, samples were limited to initial BAL samples, i.e., baseline BAL taken at the time of suspected pneumonia. For the hypothesis that specific microbiome states are indicative of clinical outcome, the null probability of a state in a pneumonia subtype being successfully, unsuccessfully, or indeterminately treated was used, i.e., a 1/3 chance of a given outcome per pneumonia state in a given disease context.

### Data and Code Availability

Sequencing data are available on NCBI SRA (pending submission). Processing and analysis scripts are available on the github repository NUSCRIPT/sumner_pneumonia_multiomics_2024.

## Data Availability

All data produced in the present study are available upon reasonable request to the authors.

## Acknowledgements

We thank Alex McFarland and Jiaxian Shen for technical expertise during initial stages. We thank our funding sources (SCRIPT NIH funding: 2U19AI135964-06; NSF GRF Grant: DGE-2234667). This research was supported in part through the computational resources and staff contributions provided by the Genomics Compute Cluster which is jointly supported by the Feinberg School of Medicine, the Center for Genetic Medicine, and Feinberg’s Department of Biochemistry and Molecular Genetics, the Office of the Provost, the Office for Research, and Northwestern Information Technology. The Genomics Compute Cluster is part of Quest, Northwestern University’s high performance computing facility, with the purpose to advance research in genomics.

## Supplemental Text

### Covariation among data types

To assess covariation between multiomic data types (Figure 1A), we employed pairwise Mantel tests on appropriate dissimilarity matrixes calculated from each omics type (Figure S2). Species-level profiles from shotgun metagenomic sequencing were compared to amplicon sequencing variant (ASV)-level and genus-level taxonomic profiles from 16S rRNA gene amplicon sequencing. Both ASV-level and genus-level taxonomic profiles from amplicon sequencing covaried with species-level abundances from shotgun metagenomic data; ASV-level data explained more variation in shotgun metagenomic taxonomic profiles than genus-level profiles as expected from the finer taxonomic resolution (Figure S2). These data support that whole genome shotgun metagenomic data reasonably capture taxonomic profiles of the lung microbiota landscape compared to deep 16S rRNA gene amplicon sequencing as a pseudo-gold standard.

Intra-omic comparison of ASV-level and genus-level 16S rRNA gene amplicons sequencing profiles are similarly significant, with 47% variance explained (Figure S2). Functional profiles from unstratified KEGG orthology (KO) term abundances were significantly correlated with species-level shotgun metagenomic profiles and genus-level amplicon profiles. RNA level features, including host transcriptomic profiles and metatranscriptomic profiles, were derived from alveolar macrophage-sorted bulk transcriptomics. These metatranscriptomic features therefore represent transcriptionally active cell-associated microbiota (e.g., internal or surface adherent). We find that covariation is low between the two RNA-based profiles and between the RNA-based and DNA-based profiles (Figure S2).

### SOFA score relationship with MDNP in NP patients

Regression analysis and Spearman rank correlation was performed between MDNP and SOFA scores to further investigate this connection (Figure S8). We observe weak, non-signficant associations between MDNP score and SOFA in patients with pneumonia independent of pneumonia-resolution.

### Bacteriophage variation associated with pneumotype classification

We identified a total of 6722 putative viral contigs across 173 of the 253 samples. Of these, 10 were identified as Complete (100%), 144 as high quality (>90%), 141 as medium quality (>50%), 5089 as low quality and 1338 could not be determined. After filtering out contigs smaller than 2.5kb and dereplication, 294 vOTUs of medium, high and complete quality were kept for downstream analysis. After removing viruses with less than 70% genome coverage and less than 10x depth, Metapop identified 79 samples containing putative viruses. Potential bacterial hosts were predicted for 158 viruses across 46 genera of host (Figure S9). The hosts with the highest number of predicted connections to vOTUs were *Streptoccocus* with 36, and *Staphylococcus* with 18. Fourteen viruses are predicted to infect more than one host, though several are still within the same genus. PERMANOVA analysis of bacteriophage community structure indicates significant associations with a few factors, notable “A: Individual” and “IB: Pneumotype” (FDR *P* < 0.01; Figure 1D).

Using bioinformatics tools for viral analysis of metagenomic assemblies, we identified putative viral contigs. Putative viral contigs co-cluster with known bacteriophage, indicating that the lung microbiome contains previously characterized bacteriophage. We observe clusters of putative bacteriophage genomes with phages of known bacterial taxa that were observed at high abundance in our samples, suggesting potential ecological interactions between bacterial and viral microbiota. Prominent bacteriophage clusters are observed between putative bacteriophages with streptococcal and staphylococcal bacteriophage, suggesting abundance and/or easily detectable phage populations associated with these genera.

## Supplemental Figures

**Figure S1.**
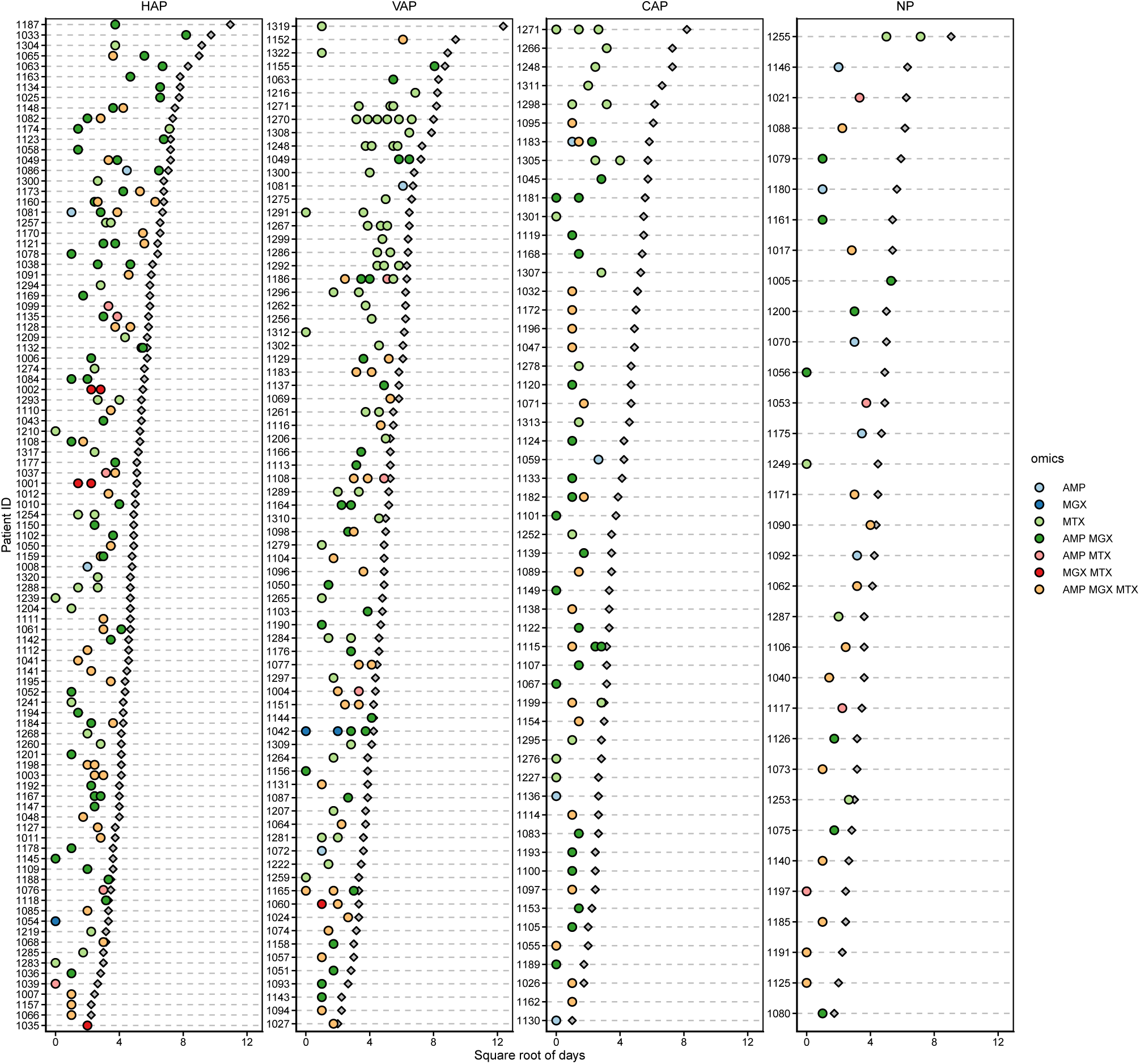
Overview of sampling per patient. Filled circles are BAL, colored by the intersection of multiomics data acquired at that time point. Grey diamonds are hospital length of stay. Note the x-axis is the square root for days.

**Figure S2.**
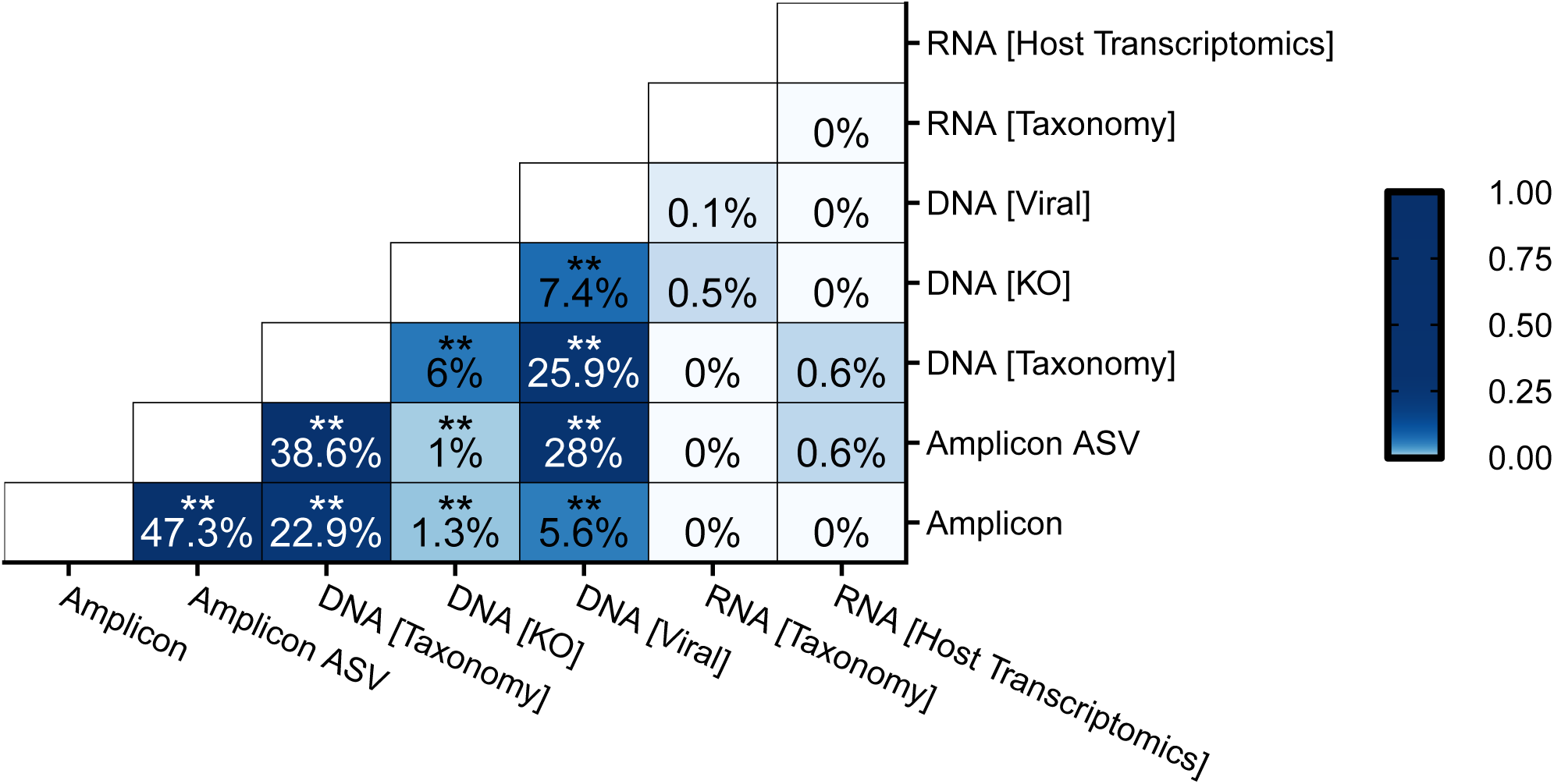
Covariation between data types tested using mantel tests. Color indicates explained variance calculated from the square of the mantel statistic. Asterisks indicate FDR *P* significance values.

**Figure S3.**
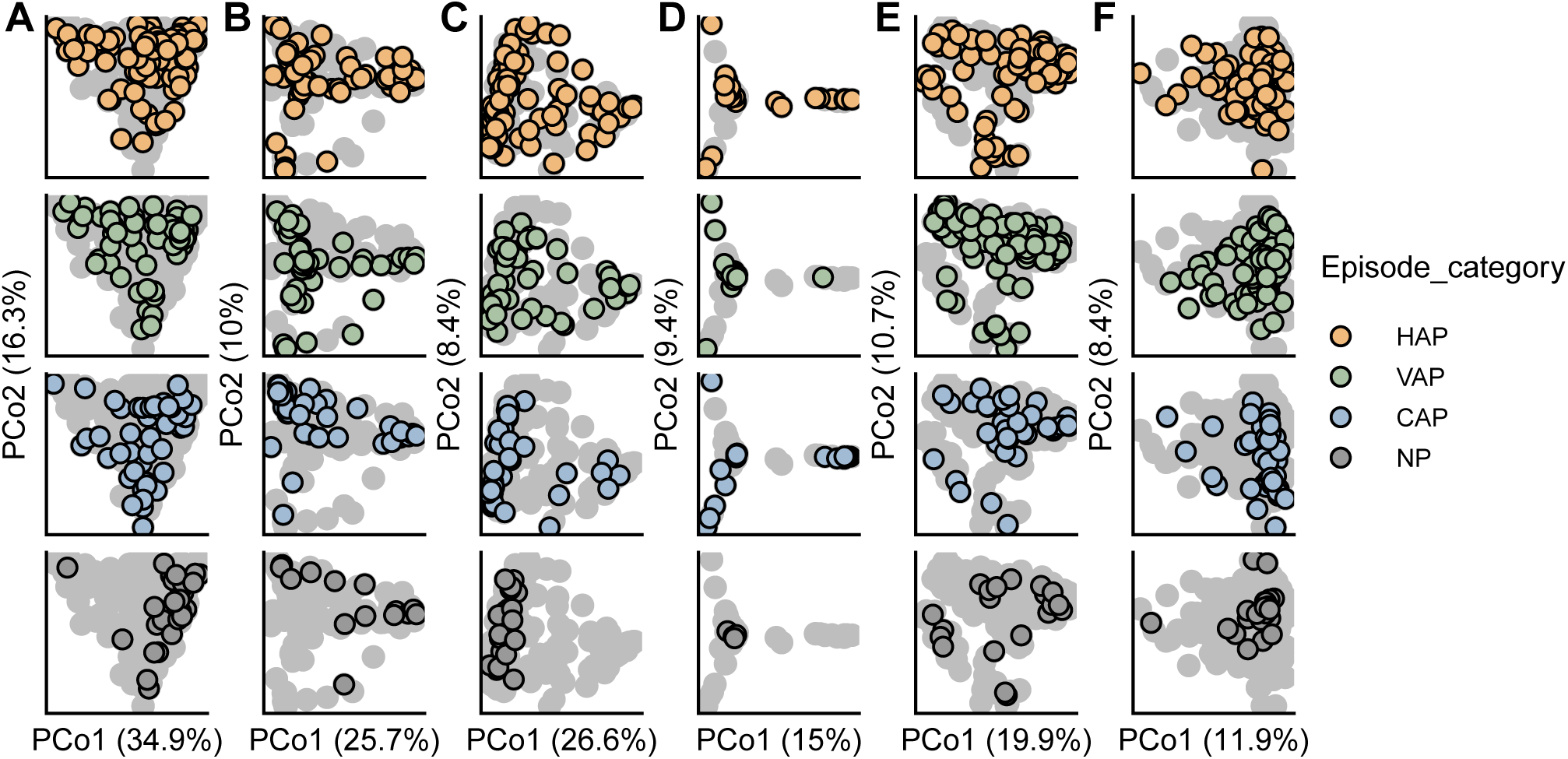
Principle coordinate analysis of multiomics data. Multiomics data include (a) taxonomic profiles from 16S rRNA gene amplicon sequencing, (b) taxonomic, (c) KEGG ortholog, and (d) bacteriophage profiles from shotgun metagenomics, (e) taxonomic profiles from metatranscriptomic, and (f) host transcriptomic profile. Weighted UniFrac used for 16S rRNA gene amplicon sequencing, and the Jaccard distance was used for bacteriophage profiles. All other multiomics dissimilarities were calculated using Bray-Curtis.

**Figure S4.**
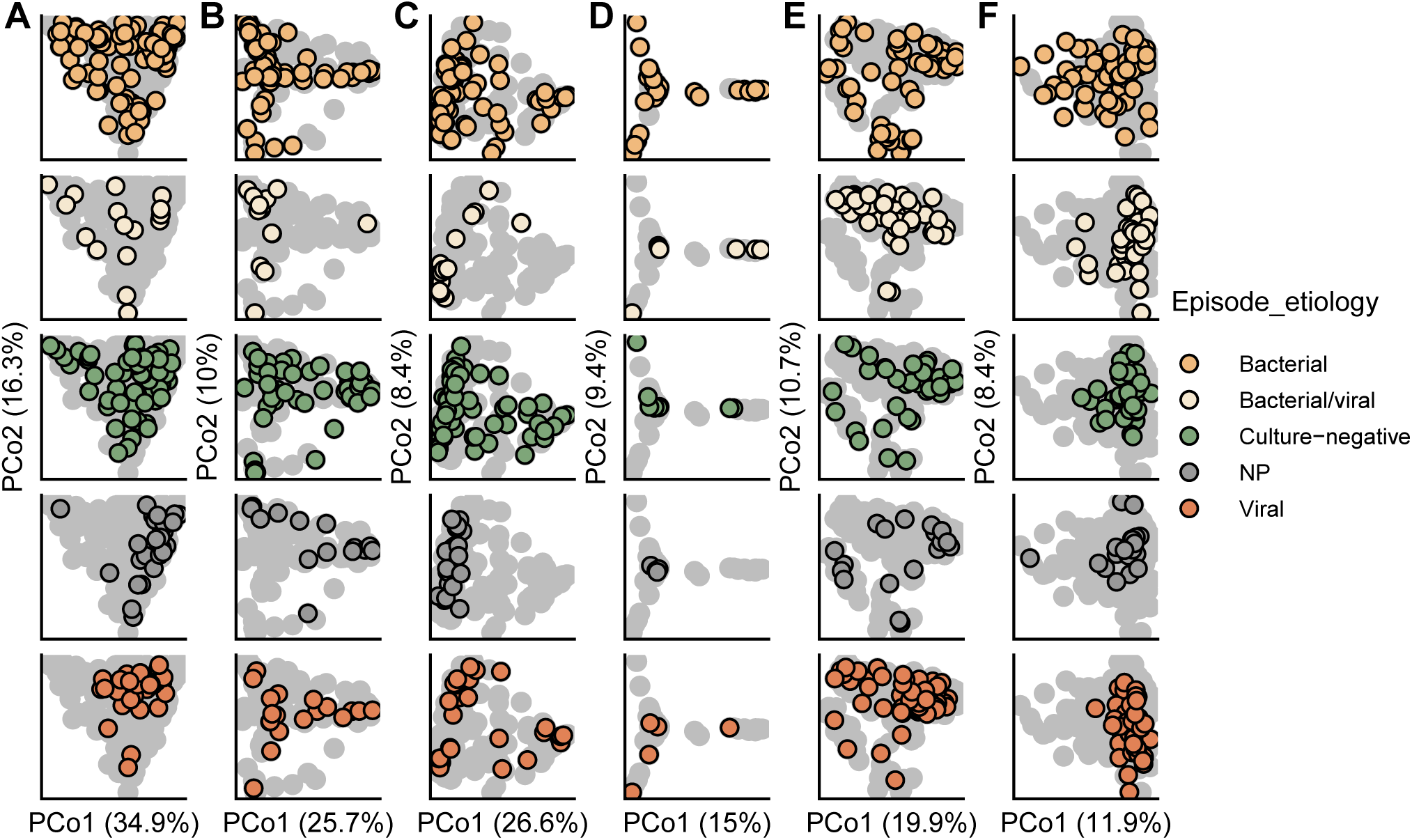
Principle coordinate analysis of multiomics data. Points colored and faceted to highlight variation in ordination by pneumonia episode etiology. Multiomics data include (a) taxonomic profiles from 16S rRNA gene amplicon sequencing, (b) taxonomic, (c) KEGG ortholog, and (d) bacteriophage profiles from shotgun metagenomics, (e) taxonomic profiles from metatranscriptomic, and (f) host transcriptomic profile. Weighted UniFrac used for 16S rRNA gene amplicon sequencing, and the Jaccard distance was used for bacteriophage profiles. All other multiomics dissimilarities were calculated using Bray-Curtis.

**Figure S5.**
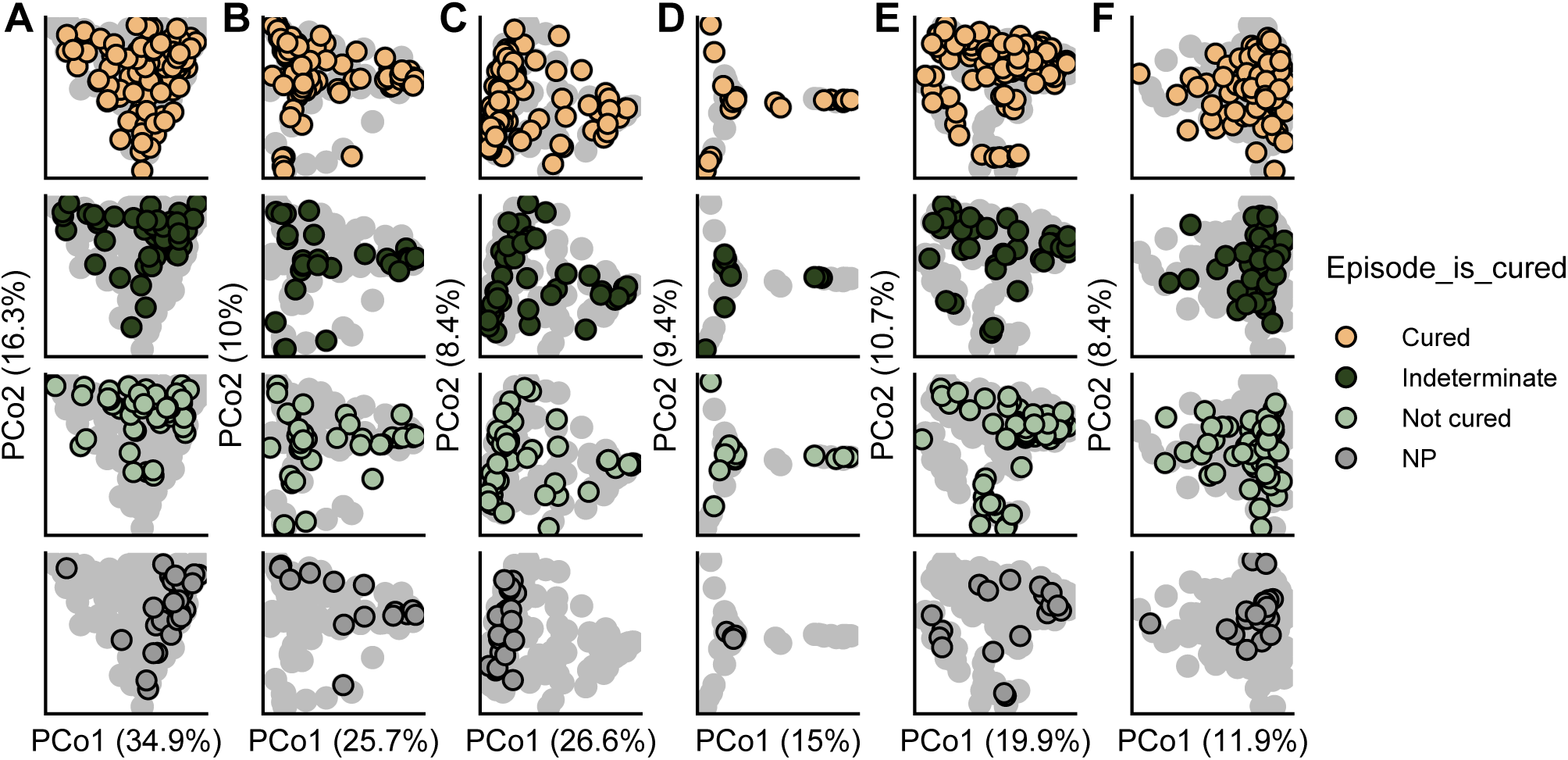
Principle coordinate analysis of multiomics data. Points colored and faceted to highlight variation in ordination by pneumonia episode clinical outcome. Multiomics data include (a) taxonomic profiles from 16S rRNA gene amplicon sequencing, (b) taxonomic, (c) KEGG ortholog, and (d) bacteriophage profiles from shotgun metagenomics, (e) taxonomic profiles from metatranscriptomic, and (f) host transcriptomic profile. Weighted UniFrac used for 16S rRNA gene amplicon sequencing, and the Jaccard distance was used for bacteriophage profiles. All other multiomics dissimilarities were calculated using Bray-Curtis.

**Figure S6.**
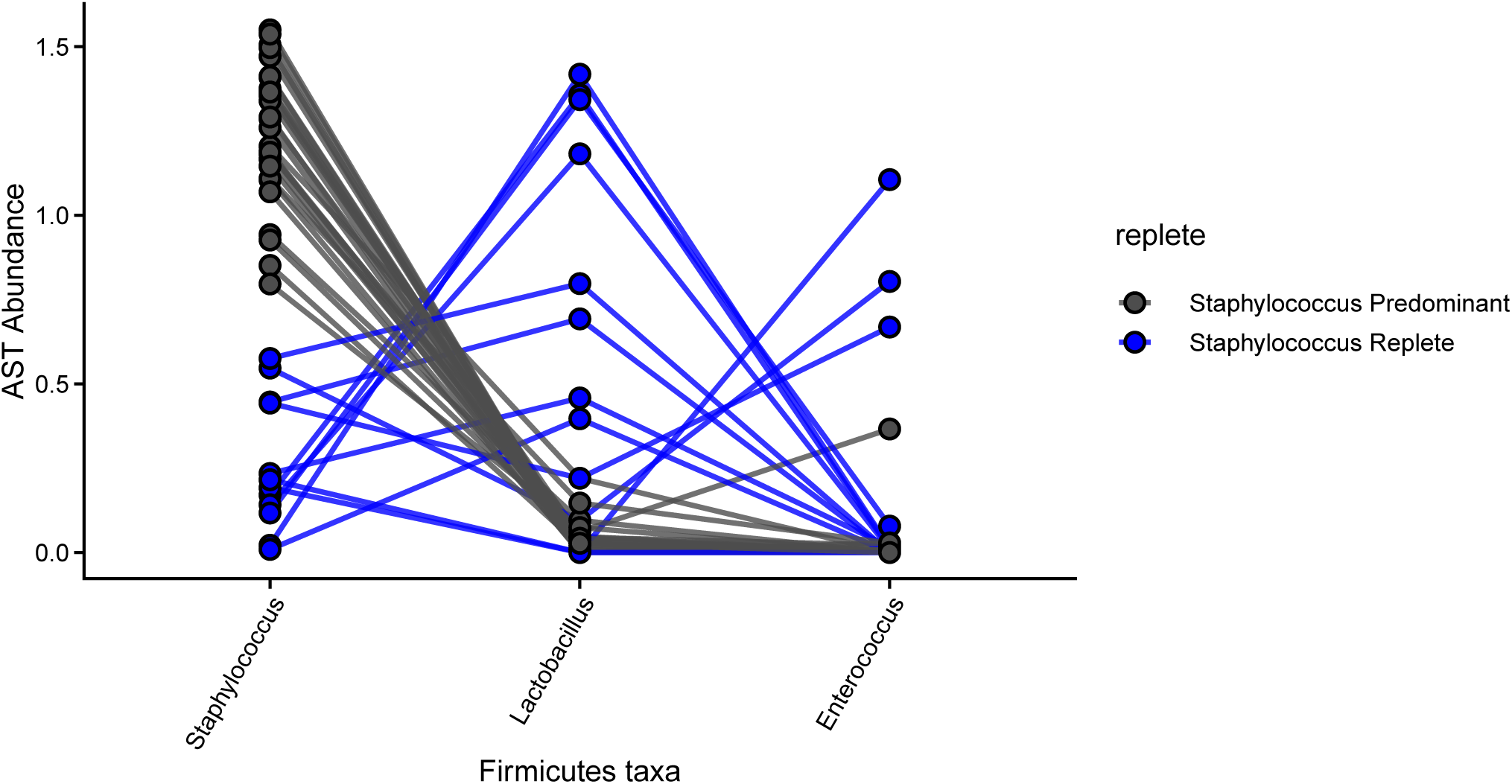
Staphylococcus is sometime replaced with other Firmicutes members in pneumotype_SP_. AST normalized relative abundance of taxa in each patient, colored by whether *Staphylococcus* is the predominant member. Samples limited to pneumotype_SP_ samples. Results indicate that *Lactobacillus* and *Enterococcus* may fulfill, at least partially, the same niche as *Staphylococcus*.

**Figure S7.**
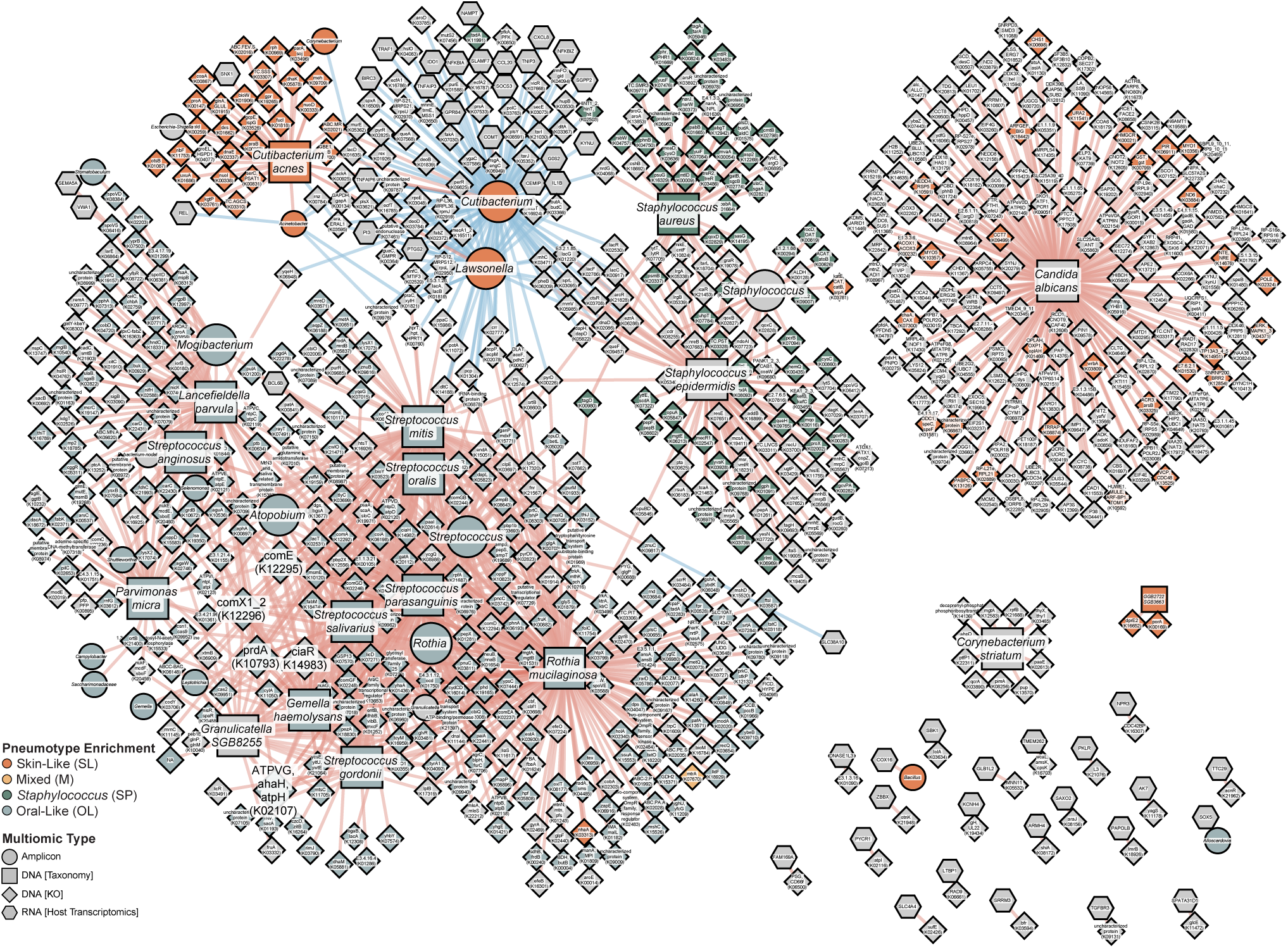
Network visualization of associated omics features identified from HALLA (see Fig. 4) but with complete labelling. Multiomics data integration includes feature profiles from four data types: shotgun metagenomic (taxonomic, functional potential), 16S rRNA gene sequencing, and macrophage-sorted bulk RNA-sequencing (host transcriptomics, metatranscriptomic). Top significant associations from each dataset comparison are visualized (FDR *P* < 0.05). Edges are associations colored by Spearman rank correlation (red for positive and blue for negative) and nodes are data features. Nodes were colored by features that were differentially over-abundant in pneumotypes; negative associations were considered to be “high” in pneumotype_SL_ as it was the baseline comparison group. Features that were high in multiple groups were kept as gray.

**Figure S8.**
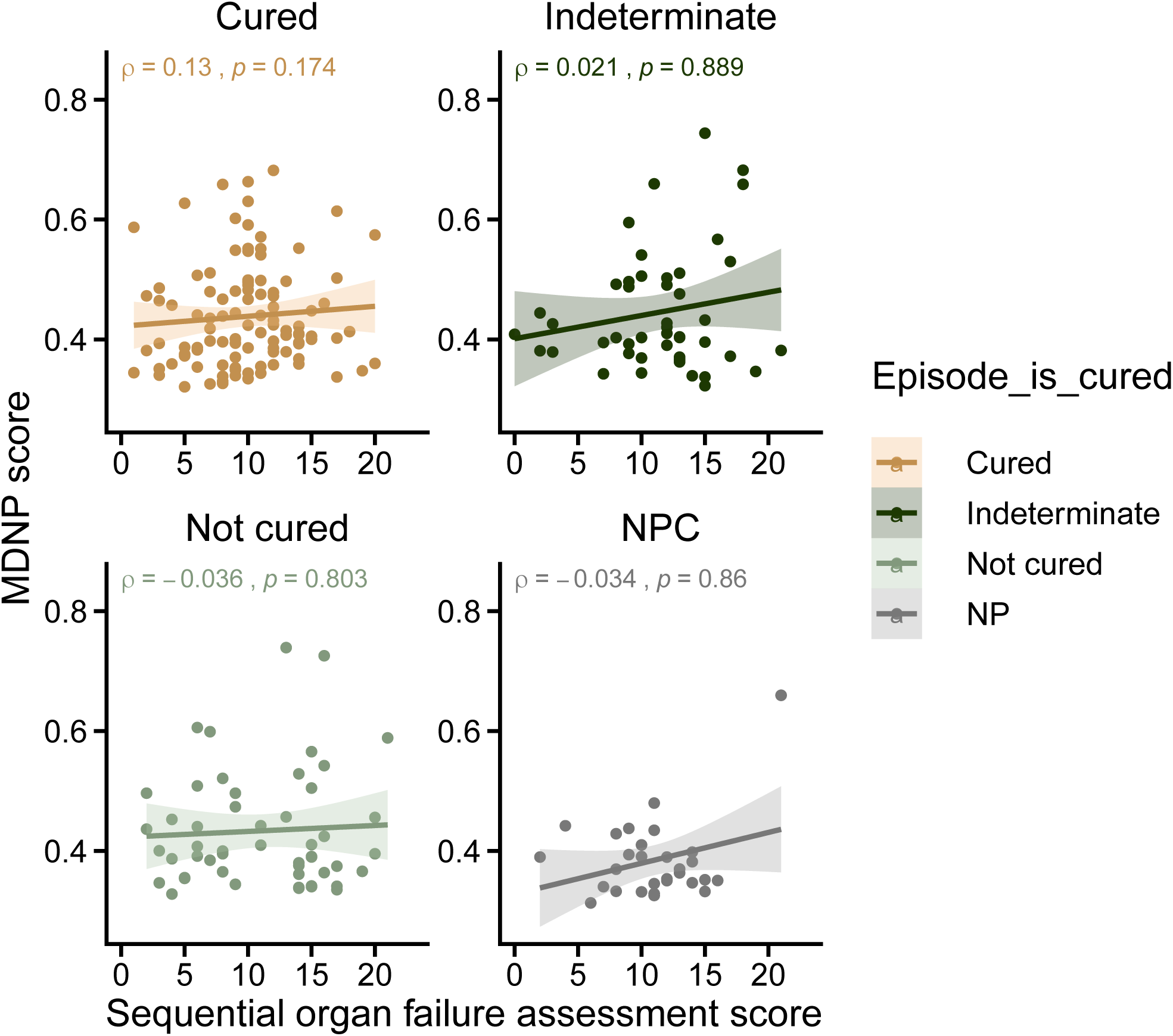
SOFA scores relationship to MDNP score. Analysis of MDNP score (mean dissimilarity to non-pneumonia) association with SOFA score. Monotonic relationship evaluated using Spearman’s rank order correlation test.

**Figure S9.**
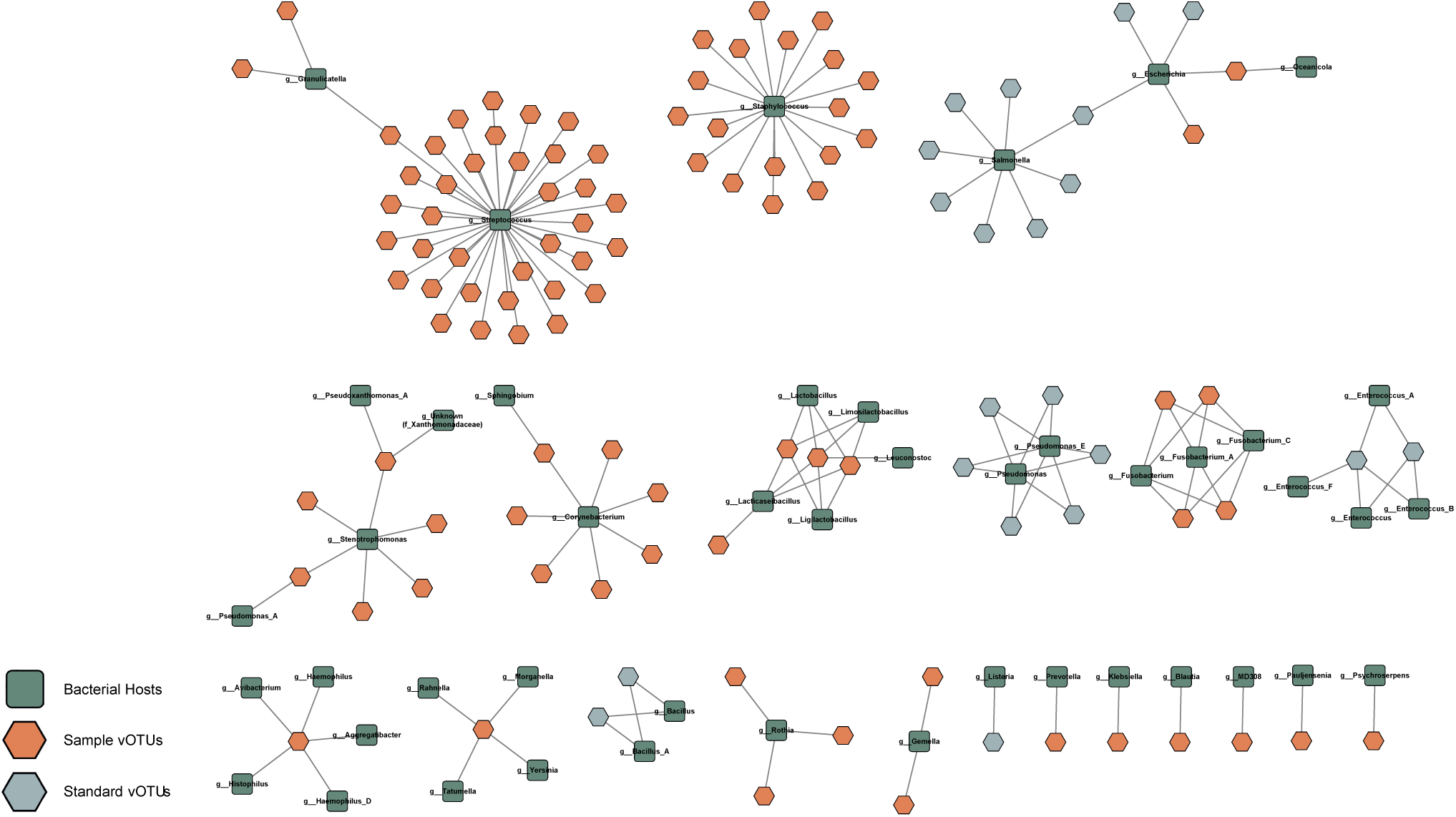
Viral operational taxonomic units (vOTUs) found in the lung. Predicted hosts (green) of vOTUs identified in BAL samples (orange) and from standards (blue). The most commonly predicted host genera are *Streptococcus* and *Staphylococcus*, both of which are found in high abundance in separate pneumotypes. Fourteen viruses identified in BAL samples are predicted to infect multiple hosts.

**Figure S10.**
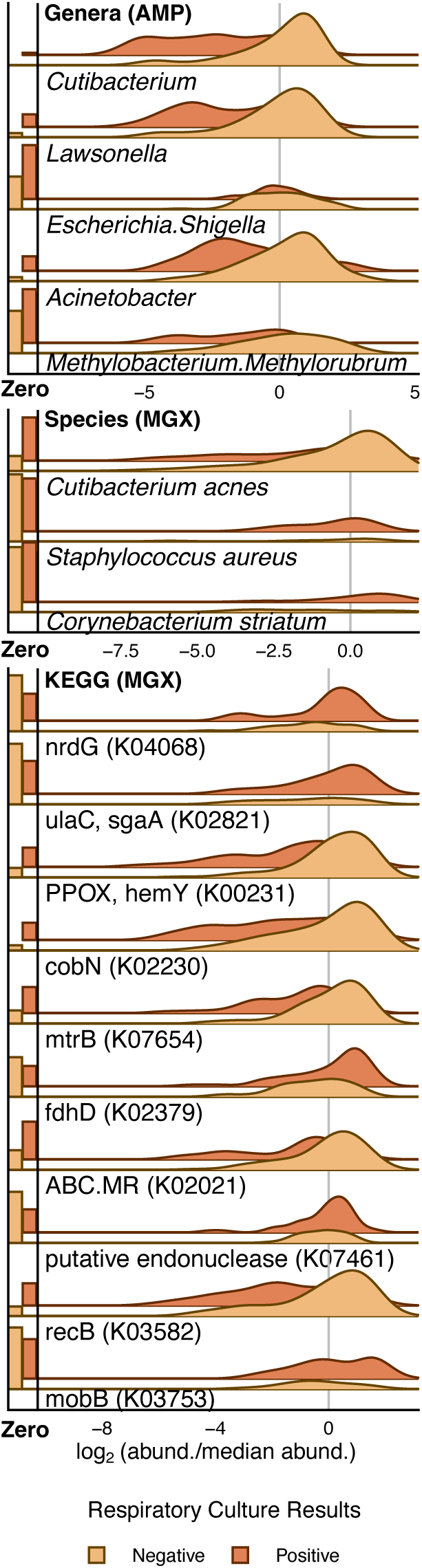
Differentially meta-omic features between respiratory culture results. Bar plots are the proportion of samples with zero-count therefore showcasing feature prevalence; bars are scaled such that touching the correspondingly colored line above indicates the feature was undetected in all samples for that group. Kernel distributions were calculated based on the subset of samples with detectable abundance after centering by the median and log_2_ transformation; heights are scaled by the proportion of detectable samples. Genes are shown with their corresponding KEGG orthology term. (* = FDR *P* < 0.05, ** = FDR *P* < 0.01, *** = FDR *P* < 0.001).

**Figure S11.**
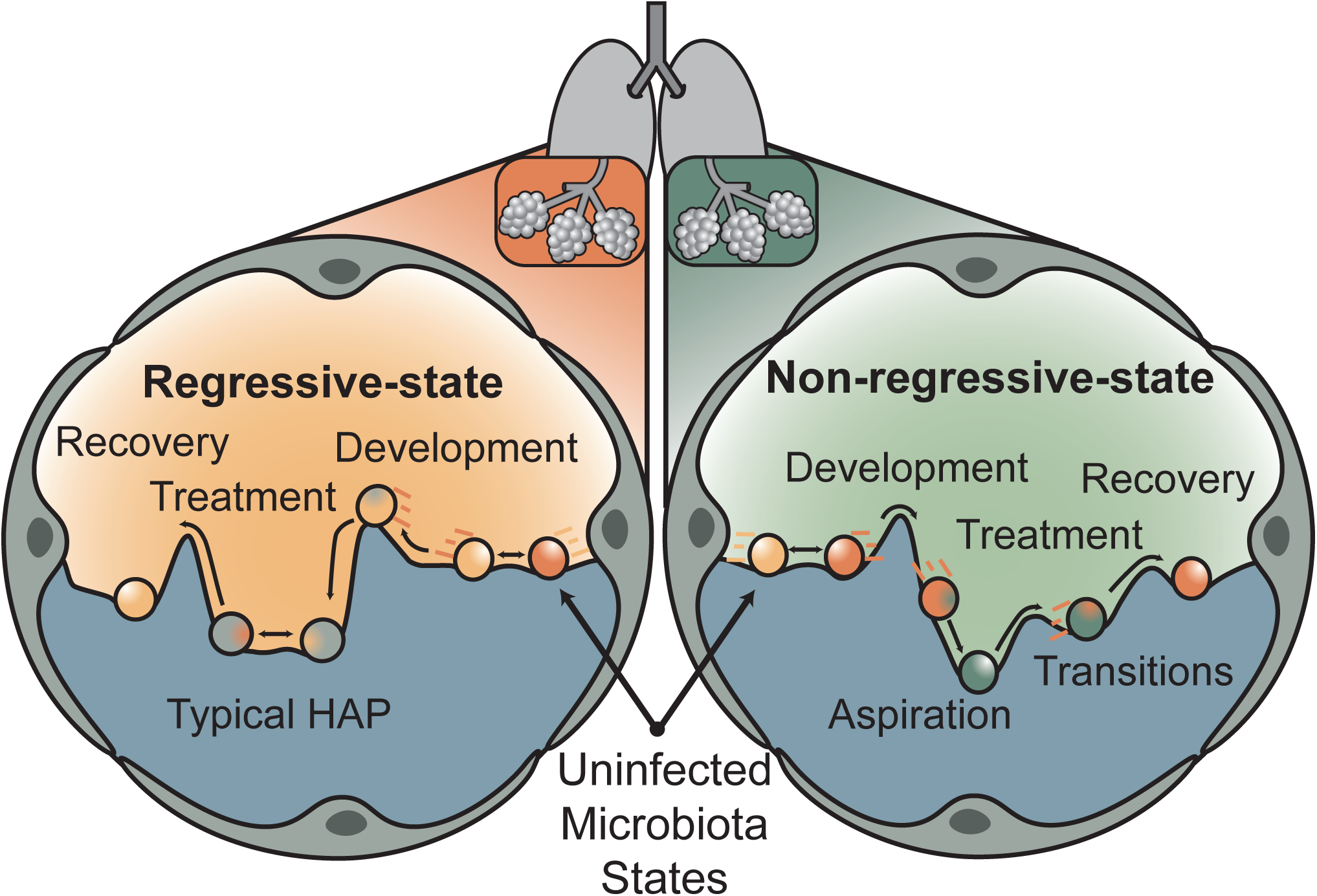
Proposed model of microbiota landscape over the course of pneumonia therapy. See discussion for details of the proposed models. Briefly, our data suggest that the lung microbiota landscape may be impacted in one of two ways over the course of pneumonia therapy depending on microbiota state stability. In a regressive-state model, microbiota state at the time of diagnosis appears globally similar to those without infection; it likely harbors non-dominant pathogens. In non-regressive dynamics, probable disruption events before diagnosis likely yield distinct, unstable microbiota states that dramatically change throughout treatment.

## Supplemental Tables

**Table S1.** Summary of case demographics. Note that pathogen etiology excludes patients who received lung transplantations.

| Category | Subcategory | n | IQR (mean) |
| --- | --- | --- | --- |
| Pneumonia Diagnosis | HAP | 97 |  |
|  | VAP | 76 |  |
|  | CAP | 54 |  |
|  | Non-pneumonia | 33 |  |
| Pathogen Etiology | Bacterial | 83 |  |
|  | Bacterial/viral | 35 |  |
|  | Viral/Etiology defined | 54 |  |
|  | Culture-negative | 63 |  |
|  | Non-pneumonia | 32 |  |
| Respiratory Culture (Bacterial) | Positive BAL | 118 |  |
|  | Negative or No Result BAL | 227 |  |
| Longitudinal Statistics | No. Patients | 62 |  |
|  | No. BAL | 156 |  |
| Quantitative Metadata | Bacterial Biomass (qPCR, Log 16S gene copies/μL) | 142 | 1.51-2.93 (2.37) |
|  | Amylase Levels (Log) | 230 | 1.20-2.53 (1.91) |
|  | Hospital LOS | 345 | 15-40 (29.2) |
|  | SOFA | 341 | 8-13 (10.6) |

